# Protein-based Diagnosis and Analysis of Co-pathologies Across Neurodegenerative Diseases: Large-Scale AI-Boosted CSF and Plasma Classification

**DOI:** 10.1101/2025.07.09.25331192

**Authors:** Ying Xu, Daniel Western, Gyujin Heo, Kwangsik Nho, Yen-Ning Huang, Shiwei Liu, Hamilton Se-Hwee Oh, Yike Chen, Jigyasha Timsina, Menghan Liu, Yinxu Tang, Katherine Gong, John Budde, Varsha Krish, Farhad Imam, Raquel Puerta Fuentes, Amanda Cano, Marta Marquie, Merce Boada, Knight Alzheimer Disease Research Center (Knight-ADRC), Dominantly Inherited Alzheimer Network (DIAN), Alzheimer Disease Neuroimaging Initiative (ADNI), ACE Alzheimer Center Barcelona (ACE), Barcelona-1, Stanford Alzheimer Disease Research Center (Stanford-ADRC), The Global Neurodegeneration Proteomics Consortium (GNPC), Pau Pastor, Agustin Ruiz, Maria Victoria Fernández, David Bennett, Tony Wyss-Coray, Andrew J Saykin, Muhammad Ali, Carlos Cruchaga

## Abstract

Neurodegenerative diseases (including Alzheimer’s disease, Parkinson’s disease, Frontotemporal dementia, and Dementia with Lewy bodies) pose diagnostic challenges due to overlapping pathology and clinical heterogeneity. We leveraged proteomic data from more than 21,000 cerebrospinal fluid and plasma samples to develop and validate explainable, boosting-based multi-disease AI classifiers. The models achieved weighted AUCs in the testing datasets of 0.97 for CSF and 0.88 for plasma, equivalent to traditional biomarkers. The model was validated with neuropathological and clinical data, confirming robust generalizability without any retraining. Using zero-shot learning, we classified disease subtypes including autosomal dominant AD and prodromal PD and clarified disease states for those with conflicting clinical information. The model also showed the ability to prioritize cognitively normal individuals at disease risk. This framework enabled the identification and quantification of continuous, individual-level disease probabilities that allow for the quantification of overlap across diseases and co-pathologies within an individual. Through this work, we establish a benchmark computational framework for enhancing diagnostic precision in NDs.

## Introduction

Dementia ranks as the seventh leading cause of mortality globally and is a major cause of disability, especially among older adults^1^. Since 2017, the World Health Organization (WHO) has emphasized the urgency of improving the early and accurate diagnosis of neurodegenerative diseases (NDs) in response to an escalating global prevalence^2^. Among the diverse spectrum of NDs, Alzheimer’s disease (AD), Parkinson’s disease (PD), Dementia with Lewy bodies (DLB), and Frontotemporal Dementia (FTD) are some of the most common^3^. Notably, AD alone is responsible for approximately 60-80% of dementia cases^4^, while PD is widely recognized as the second most common progressive neurodegenerative condition^5^. DLB and FTD, while less common, represent essential considerations for clinicians and researchers alike because of their distinctive pathology, clinical presentation, and growing recognition as contributors to the dementia burden. Although these diseases are characterized by neuropathological changes (amyloid, tau, α-synuclein, TDP-43, and fused-in-sarcoma (FUS), there is overlap in these hallmarks across diseases.

Currently, there are highly accurate cerebrospinal fluid (CSF) and plasma AD biomarkers, including amyloid beta 42 (Aβ42)^6^, phosphorylated tau at threonine-217 (pTau217), and pTau181^7^. Neurofilament light chain (NFL) and glial fibrillary acidic protein (GFAP) have been proposed as potential biomarkers for DLB or FTD, but those are more likely to be general neurodegenerative markers rather than disease-specific. Novel assays for synuclein are being developed that show promising results for PD, but there are currently no well-validated biomarkers for three of the four most common neurodegenerative diseases. In addition, even the existing biomarkers do not enable the identification and quantification of overlapping pathology within an individual, as they are typically developed to ensure disease specificity although existence of multiple pathologies is widespread based on postmortem^8^ neuropathological evidence.

Over the last decade, proteomic-based studies have emerged as a powerful approach in the field of dementia research^9–13^, supplementing or even surpassing genomics-based strategies in certain contexts, especially for prediction and identifying pathways relevant to the disease^14^.

Proteomic analyses offer a direct window into the functional components in dementia development and pathogenesis, implicating neuronal integrity, synaptic transmission, and neuroinflammatory processes characteristic of dementia^15^. Several large-scale studies of these diseases have mapped protein expression patterns in cerebrospinal fluid (CSF) and plasma^9,12,16–18^, demonstrating that proteomic biomarkers have potential for early and precise disease detection, mechanistic insights, and tailoring of therapeutic interventions. However, few proteomic analyses have been performed in DLB and/or FTD, while those that have been published are limited to relatively small datasets^19–22^.

Additionally, none of these studies completely capture the inherent complexity and heterogeneity across neurodegenerative disorders. Many individuals with neurological decline exhibit overlapping clinical features or biomarker signatures, complicating the accuracy of traditional case vs. control analyses. There is a need to generate robust CSF and plasma predictive models that can accurately classify individuals using a paradigm that improves upon simple binary case vs control models through differentiation between controls and multiple forms of disease at once.

Artificial Intelligence (AI) methods have begun to improve dementia diagnosis by combining heterogeneous multi-modal datasets^23^, including electronic health records, medical imaging, cognitive profiles, and single-analyte biomarkers. However, most existing tools remain binary (disease versus control)^24^ and seldom address the biological overlap across dementia types that characterizes neurodegeneration. In practice, clinicians must discriminate not only between normal aging and disease-associated changes but also between different forms of dementia, and this is especially challenging in the earlier prodromal stages of the diseases where clinical changes and even well validated biomarkers may be less informative. In top of this, many patients can present co-pathologies. For instance, around 30% of the AD cases, present α-synuclein pathology and a large proportion of PD cases also present tau or Aβ pathology, making diagnosis and treatment even more challenging. Algorithms that can assign continuous probabilities across multiple diagnoses, rather than a single label, are therefore essential to detecting mixed pathology, determining those at high risk of dementia development, and providing a more granular picture of ND risk.

Among the diverse family of AI-based classification algorithms, tree-based models achieve state-of-the-art performance on structured omics data while maintaining transparency^25^, a key feature of an explainable model. Their additive tree structure allows straightforward extraction of global and class-specific feature-importance scores, mitigating the “black-box” concern often associated with AI approaches such as deep neural networks^26^. A recent advanced approach called zero-shot^27^ inference borrowed from natural-language processing further enables such boosted models to assign meaningful probabilities to phenotypes that were not directly included in samples from the training distribution. This ability offers a route to classify rare or emerging disease types into their most similar common alternatives, which may enable improved treatment for individuals with those types.

To increase the interpretability of these methods, AI-based computational modeling must be anchored to biology^28^. A clinically meaningful classifier should show concordance between predicted disease probability and disease-relevant markers, such as increasing AD likelihood with rising neuritic plaque load or Braak neurofibrillary tangle stage and consistently low probabilities in pathologically confirmed controls. The model-assigned AD probability should correlate inversely with scores on cognitive examinations, including the Mini-Mental State Examination (MMSE) and episodic memory tests, reflecting the expected coupling between pathological burden and cognitive decline. Consistency between predicted disease states and biological outcomes provides additional evidence for clinical utility beyond typical discrimination metrics (AUC, F1 score) alone, ensuring that the model captures genuine biologically relevant decline.

In the present study, we leveraged large CSF and plasma proteomic datasets to develop and validate an explainable boosting-based multi-class classifier that encompasses the most common NDs. Our approach relied exclusively on proteomic features, foregoing any demographic or clinical variables, to underscore the predictive value of biomarker profiles alone. We validated the models across a wide range of cohorts to ensure reproducibility across clinical environments and populations. The model performance was benchmarked in independent dataset against neuropathological, clinical phenotypes and well validated imaging phenotypes without data relabeling, retraining or imputation. Finally, the model was tested against seen disease types (Zero-shot), such as such as autosomal dominant AD (ADAD) and Parkinson’s disease with dementia (PDD), and individuals without clear clinical diagnosis.

## Results

### Study Design

We utilized a multi-stage approach for the development of our AI classifier, where we first trained our model using internal CSF & plasma proteomic data from five neurodegenerative disease categories (**Fig. 1, Supplementary Table 1A**). The internal dataset includes proteomics from 4,677 samples from seven cohorts (Knight Alzheimer’s Disease Research Center (ADRC), Stanford ADRC, Washington University Movement Disorder Clinic (MDC), Barcelona-1, ACE, ADNI, and PPMI). This included 2,230 samples from individuals that were assigned to one of our core five diagnoses (665 healthy controls (negative for AD-relevant biomarkers^29,30^), 744 AD (biomarker positive), 738 PD, 46 FTD, and 37 DLB cases). The remaining 2,447 samples were obtained from individuals with a range of diagnoses that were used for further benchmarking of the model or for classification purposes. These included individuals with ADAD, prodromal PD, or PDD, as well as people with evidence of dementia but unclear clinical diagnosis (other or OT; **Supplementary Table 1B)**. Furthermore, this subset of samples included cognitively normal individuals with AD pathology (preclinical AD), or individuals with preclinical signs of PD (prodromal PD). In parallel, we assembled a plasma dataset of 4,750 samples from three cohorts (Knight ADRC, Stanford ADRC, and MDC), comprising 1,282 AD biomarker negative healthy controls, 865 AD (biomarker positive), 687 PD, 44 FTD, and 122 DLB cases. Like in CSF, 1,750 samples were obtained from individuals without clean grouping into those categories that were used for benchmarking and further testing the model.

**Figure 1.**
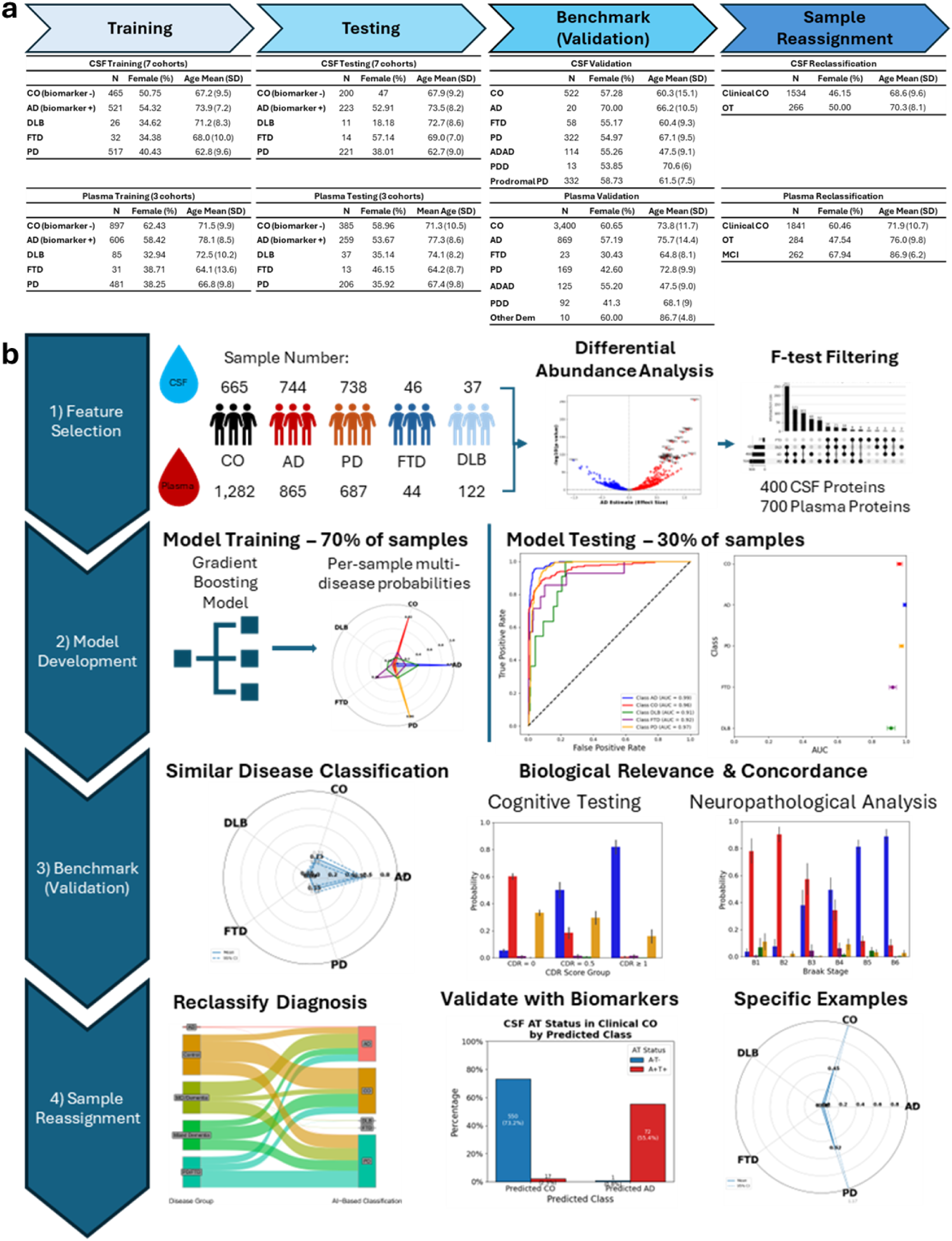
Study Design. A. Sample Demographics. CSF samples from the training & testing datasets were obtained from the Knight ADRC, ADNI, Stanford ADRC, MDC, Barcelona-1, Fundacio ACE, and PPMI. Plasma samples from the training and testing datasets were obtained from the Knight ADRC, Stanford ADRC, and MDC. Samples used for validation were accessed through DIAN, ROSMAP, the I-ADRC, and the GNPC. Samples used for reclassification were from the Knight-ADRC & ROSMAP. **B. Workflow Steps.** Proteomic data (SOMAscan 5k or 7k) from five main diagnostic categories, two tissues, and multiple cohorts (Knight-ADRC, ADNI, Stanford ADRC, Barcelona-1, Fundacio ACE, MDC, and PPMI) was used to identify differentially abundant proteins in each disease compared to controls. Those proteins were then filtered based on their importance and were used to build a tree-based gradient boosting AI disease classifier (LightGBM) using a training-testing paradigm. Using independent external data from ROSMAP and the I-ADRC along with related disease categories from internal data, validation of the model performance was done using relevant cognitive and neuropathological phenotypes. Finally, the classifier was used to clarify diagnoses for controls with biomarker evidence of disease and individuals with unclear disease etiology and to investigate specific samples with characteristics of interest, Control; AD, Alzheimer’s Disease; PD, Parkinson’s Disease; FTD, Frontotemporal Dementia; DLB, Dementia with Lewy Bodies, ADAD, autosomal dominant Alzheimer’s disease; PDD: Prodromal PD; OT, other dementia; MCI, mild cognitive impairment.

We leveraged CSF and plasma samples from the internal dataset to perform a comprehensive protein feature selection (**Fig. 1**). We then used those features to build an AI classifier using a robust boosting-based machine learning model (LightGBM^31^). We trained and tested the models in each fluid using a 70/30 internal split and validated the performance by applying the model to related but independent disease outcomes (autosomal dominant AD (ADAD), prodromal PD, and PD with dementia (PDD)) using zero-shot AI principles. We evaluated the performance of the model by applying it without retraining on external plasma datasets from the Religious Orders Study – Memory and Aging Project (ROSMAP) and the Indiana ADRC (I-ADRC). Once we validated the AI classifier across a range of conditions and cohorts, we sought to use the model to prioritize controls at risk of ND development, including cognitively normal samples with evidence of AD biomarkers, and compared the model probabilities to neuropathology data. We also applied the model to samples from subjects with unclear diagnosis (“other” or OT) to refine their diagnoses. We further analyzed external CSF & plasma data from the Global Neurodegeneration Proteomics Consortium (GNPC) to assess performance in a broad-scale proteomic dataset of diverse, heterogenous sample origin (**Supplementary Table 1B**). Using biomarker, cognitive exam, and neuropathological data from the Knight-ADRC, ROSMAP, and GNPC, we assessed the concordance between the AI model’s results and disease-relevant biology, ensuring our findings were grounded in relevant phenotypes. Finally, to emphasize the clinical relevance of the AI-based classifier, we highlighted specific examples where the model was supported by robust biological evidence, both within concordant diagnoses and classifications and in instances of reclassification where we observed evidence of improved diagnostic accuracy using our model.

### AI-based Multi-class ND Classifier

#### Disease-associated Analyte Selection

We first sought to identify specific analytes upon which we could base the development of our classifier model. To reduce the feature space, we first performed linear regression analyses in each fluid to identify proteins associated with four well-known neurodegenerative diseases (NDs), AD, PD, FTD, and DLB, by comparing normalized protein levels in each disease group against healthy controls. All models were adjusted for age, sex, and the first two protein principal components (PCs) to control for confounding factors **(Supplementary Fig. 1)**.

Because some CSF cohorts were analyzed using different versions of the SomaScan platform (SomaScan 7k & 5k), we included only analytes that were present in both versions. Among 3,622 common analytes **(Supplementary Fig. 2)**, we identified 2,020 nominally associated with AD, 1,809 with PD, 2,024 with FTD, and 1,281 with DLB in CSF (**Supplementary Tables 2 & 3, Supplementary Fig. 3**). From the 6,607-analyte plasma panel, we detected 1,719 proteins nominally associated with AD, 2,394 with PD, 522 with FTD, and 953 with DLB **(Supplementary Table 4, Supplementary Fig. 4)**. Taking the union of these disease-related analytes yielded 3,020 disease-associated features in CSF and 3,607 in plasma.

To further refine these feature sets, we applied an F-test-based approach, which quantifies how much of each analyte’s total variance is explained by diagnostic class relative to within-class noise. This step reduced the features to 400 analytes in CSF and 700 in plasma (**Fig. 2A&B**). Within the 400 CSF features, 18 analytes were specific to AD (such as GDA, PDE6D, FN1), seven to PD (SEMA4B, TNFSF8), and three to FTD (VSIG2, GLIPR1, IGFBP4), whereas 147 (AK2, EIF3G, MDH1) were shared by all diseases (**Fig. 2C&E**). Shared molecular signatures were observed across multiple diseases, including MSMP, NPNT, and CCDC80 in CSF for both PD and DLB, TNFSF12, UCHL1, and FLT1 in CSF for both AD and FTD, and FLII, CCDC50, RNF215 in CSF for both AD and PD. Among the 700 plasma analytes, 68 were specific to AD (NEFL, MSMB), 62 to PD (CBFB, STX12), four to FTD (HS3ST5, SNRPD3, BDP1, ACTN2), and two to DLB (PRXL2A, IL17F), with just 26 (ALDH1A1, SPP1) commonly expressed among all four NDs (**Fig 2D&F, Supplementary Fig. 4**). Many proteins were shared between two diseases in plasma, including ART3 and ACHE for AD and DLB and NPDC1 and COL28A1 for PD and DLB.

**Figure 2.**
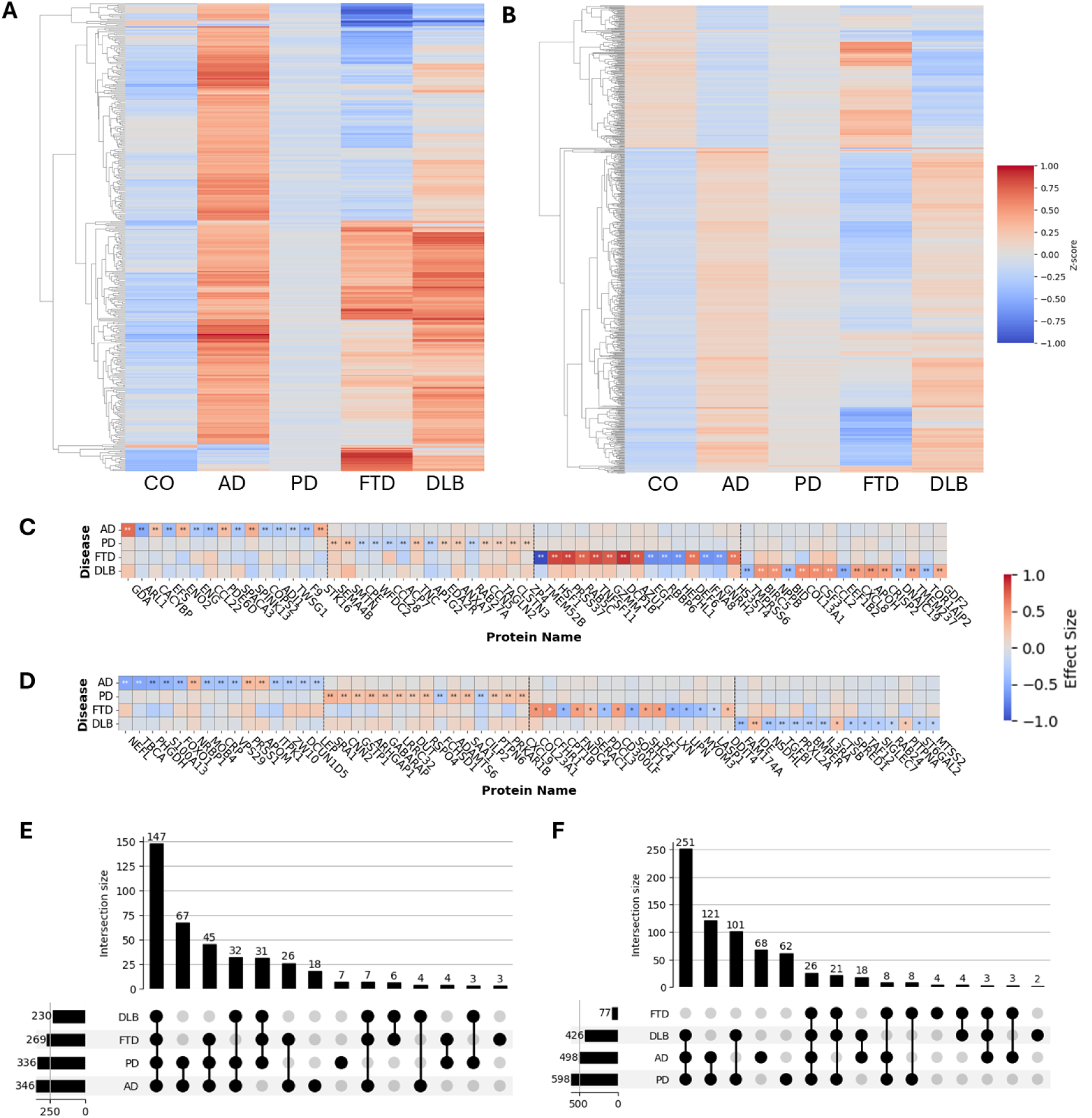
Differential Protein Expression in CSF and Plasma Across Neurodegenerative Diseases. (A, B) Heat Maps of Averaged Protein Expression in CSF and Plasma. Average z-scored expression profiles of 400 cerebrospinal fluid (CSF) analytes (A) and 700 plasma analytes (B) used for downstream classification, spanning AD, PD, FTD and DLB. Rows represent individual analytes, while columns represent mean expression values for each disease category. Warmer colors (red) indicate higher relative expression levels, and cooler colors (blue) indicate lower relative expression. **(C, D) Disease-Specific Analyte Heat Maps (Top 15 per Disease).** In C (CSF) and D (Plasma), the top 15 nominally significant proteins for each disease are arranged by effect size, yielding a combined total of 60 analytes. Color intensity reflects the absolute value of the effect size, with red indicating higher expression or stronger positive associations and blue indicating lower expression or negative associations. Asterisks highlight significant changes (* p < 0.05; ** FDR < 0.05), underscoring potential biomarker candidates for differential diagnosis. **(E, F) UpSet plots of F-test–selected analytes.** Panels E (CSF) and F (plasma) display the intersections among the final feature sets obtained after variance filtering and F-test ranking. The CSF panel summarizes 400 retained analytes, whereas the plasma panel summarizes 700 analytes. Bars above each plot indicate the number of analytes shared by the corresponding disease combinations, highlighting both disorder specific markers and analytes common to multiple neurodegenerative conditions.

#### Multi-class Classification modeling and performance

Following the identification of 400 CSF and 700 plasma analytes with relevance to neurodegenerative disease, we employed explainable AI-boosted models (LightGBM^31^) using 70% of the CSF and plasma proteomic data (**Table 1**) to build classifiers that could distinguish between healthy controls (CO, AD biomarker negative), AD (AD biomarker positive), PD, FTD, and DLB cases exclusively from proteomic data. Both models showed excellent performance, with AUCs > 0.96 and accuracies of > 94% for all the diseases **(Supplementary Fig. 5, Supplementary Tables 5 & 6)**. Using Shapley Additive exPlanations (SHAP), we investigated the proteins that most strongly contributed to the model probabilities in each fluid. Based on the SHAP analyses, we identified 14-3-3 protein gamma (YWHAG)^9,32^ in CSF and ACHE in plasma^33^ as proteins that strongly differentiated between AD and CO. CSF SETMAR, on the other hand, can help to distinguish FTD and PD (**Supplementary Fig. 6 & 7, Supplementary Table 7**), and ARRDC3 stratifies PD from other diseases (**Supplementary Fig. 6 & 7, Supplementary Table 7).**

In the CSF testing dataset (30% of our samples), the classifier achieved a macro AUC of 0.95, a weighted AUC of 0.97, and an overall accuracy of 87.89% (**Fig. 3a**). The model performance was robust for AD (precision=0.91, recall=0.96, F1-Score=0.93; AUC=0.99), CO (precision=0.91, recall=0.83, F1-Score= 0.86; AUC=0.96), and PD (precision=0.85, recall=0.92, F1-Score=0.89; AUC=0.97). FTD (n = 14) and DLB (n = 11) were limited to relatively small samples sizes and yielded F1 scores of 0.40 and 0.15, respectively **(Supplementary Table 9A & 9B, Supplementary Fig. 8).** Overall, the model assigned correct classes with high predictive probabilities. Biomarker-confirmed AD cases had an average 94% probability of being AD while PD cases had 90% mean probability for PD (**Fig. 3A, Radar Plot**). Additionally, 95.5% of biomarker-confirmed AD cases were correctly classified as AD and 92.3% of PD cases were classified as PD, exemplifying the accuracy for the common classes. Importantly, while we had access to biomarker information for AD that increased confidence in the original assignment of our controls and AD cases used for training & testing, we depended strictly on clinical diagnosis for the other three categories, which may affect model training and subsequent accuracy in these categories. To date, no highly accurate biomarkers are available for PD, FTD, or DLB. Additionally, FTD & DLB often show substantial symptomatic overlap with AD and other forms of dementia, making their diagnosis more difficult and potentially affecting our ability to classify those diseases.

**Figure 3.**
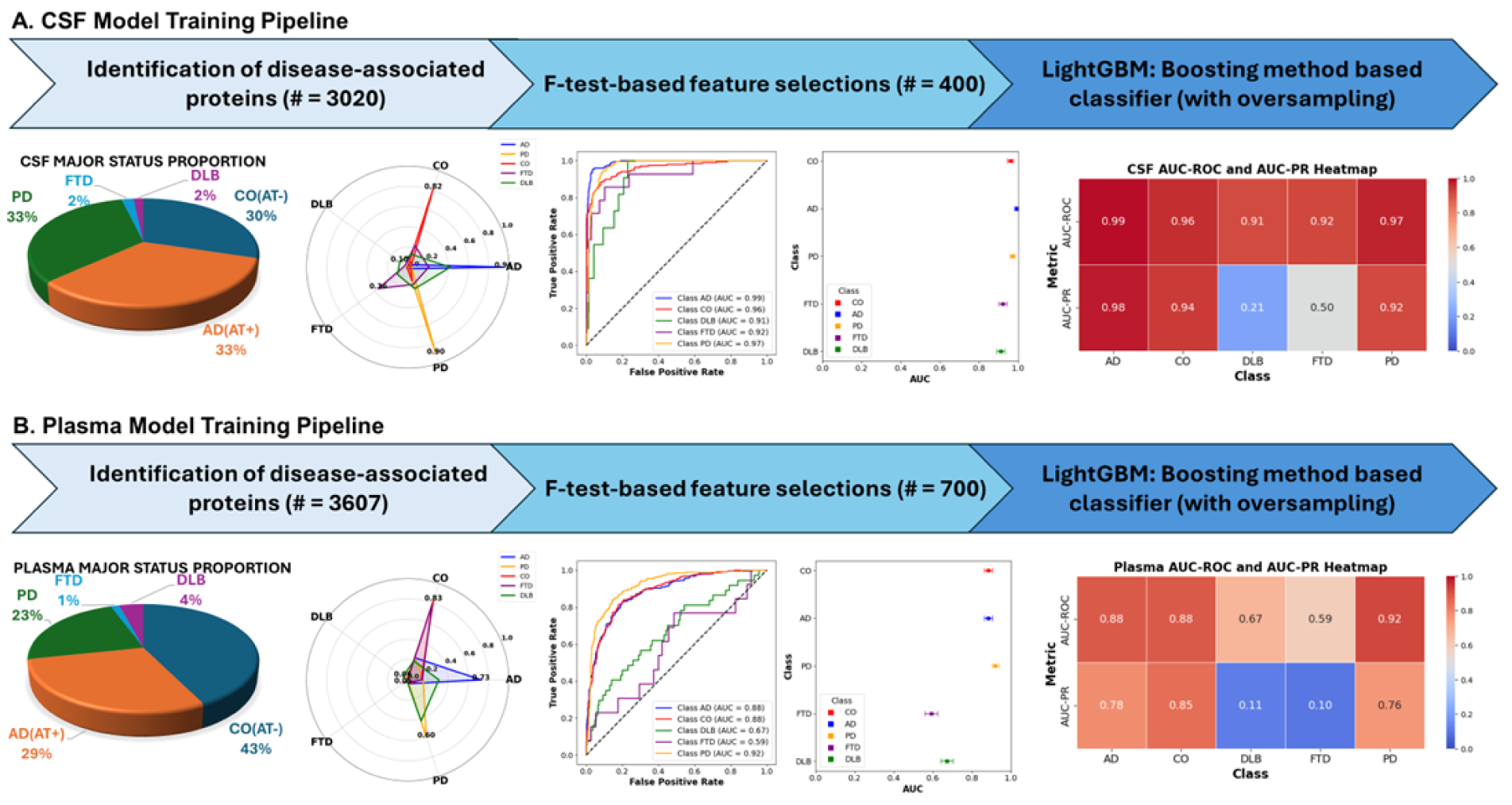
CSF and Plasma Multi-panel Performance. (A) CSF Classifier Performance. The upper flow chat shows the schematic overview of the cerebrospinal fluid (CSF) workflow, beginning with differential abundance analysis and subsequent F-test-based feature selection to narrow the set to 400 top-ranked analytes. A LightGBM model with SMOTETomek oversampling was then trained for multi-class discrimination among CO, AD, PD, FTD, and DLB. The lower panels show CSF internal 30% hold-out testing performance. Panel 1: clinical status for samples included in the CSF testing dataset. Panel 2: Radar plot depicting the probability distributions (mean and 95% CI) for each of the five major status categories. Panel 3: AUC curves for classification accuracy for each disease alongside the model’s overall weighted AUC (0.97) and macro AUC (0.95).Panel 4: Whisker plot for AUCs for all five classes with 95% confidence interval (CI). Panel 5: Heatmap summarizes both AUC-ROC and AUC-PR for the five classes. **(B) Plasma Classifier Performance.** Again, the upper flow chart showed the schematic overview of the plasma workflow, initiated with differential abundance analysis (3,607 disease-associated analytes) followed by variance-and F-test-based feature selection to yield 700 top-ranked analytes. A LightGBM model with SMOTETomek oversampling was then trained to classify CO, AD, PD, FTD, and DLB. The lower multi-panels showed plasma internal 30% hold-out testing performance. Panel 1: clinical status for samples included in the plasma testing dataset. Panel 2: Radar plot depicts probability distributions (mean and 95% CI) for each of the five major status categories. Panel 3: AUC curves for classification accuracy for each disease alongside the model’s overall weighted AUC of 0.88 (macro AUC 0.79). Panel 4: Whisker plot for AUCs for all five classes with 95% CI. Panel 5: Heatmap again shows AUC-ROC and AUC-PR for the five classes.

In plasma, the same methodological pipeline yielded a weighted AUC of 0.88 and a macro AUC of 0.79 (overall accuracy 73%, **Fig. 3b**). PD demonstrated the highest performance for a single status (AUC = 0.92; precision=0.81, recall=0.64), suggesting that PD-related proteomic changes may be captured well in plasma, consistent with PD having the most differentially abundant proteins in plasma. AD (AUC = 0.88) and CO (AUC = 0.88) were close behind, while FTD and DLB showed lower performance (FTD AUC = 0.59, DLB AUC = 0.67, **Supplementary Table 10A & 10B**), potentially due to a combination of factors including sample size limitations, clinical diagnosis mistakes and overlap between these diseases and the more common NDDs. Although plasma assays may not capture ND-relevant molecular changes to the same degree as CSF, these results highlight plasma proteomics as a viable, minimally invasive platform for biomarker-driven disease classification.

To demonstrate the effectiveness of our model compared to gold-standard biomarkers for AD, we calculated the AUCs for models based on either plasma pTau217 or amyloid PET imaging, adjusted for age and sex using the individuals from both the CSF and plasma testing datasets. The pTau217-based models yielded AUCs of 0.88 (CSF data) and 0.91 (plasma data; **Supplementary Tables 11**). Models based on amyloid PET imaging attained AUCs of 0.94 (CSF) and 0.88 (plasma). Our proteomics-driven LightGBM classifier achieved an AUC of 0.97 in CSF and 0.88 in plasma, showing performance comparable to plasma pTau217 and amyloid PET imaging while simultaneously distinguishing AD, PD, DLB, FTD, and CO. Thus, our model provides competitive single-disease accuracy together with cross-disease differentiation. The slightly higher AUCs observed here for the AD biomarkers relative to earlier reports likely reflect our filtering of AD cases and controls using biomarker status, which tightens the correlation between biomarker status and clinical diagnosis. When applying the pTau217 or amyloid PET models to clinical status (without biomarker filtering), performance was comparable to other estimates^34^ as expected (AUC using pTau217 for clinical AD vs CO = 0.78 vs 0.80 in similar work).

In summary, we developed CSF & plasma boosting tree-based models that accurately classify samples across a range of neurodegenerative diseases, illustrating the potential of proteomics-driven multi-class classifiers to robustly identify a spectrum of neurodegenerative conditions, even without imputation or demographic information.

### Clinical Relevance at Scale

#### Validation of model based on neuropathology and clinical data

To evaluate the generalizability of the classifier beyond the cohorts and clinical diagnoses used for model development, we used the zero-shot AI approach to apply it, without retraining or imputation, to diagnostic categories that are similar to one of the original five categories. In CSF, we applied the model to samples with autosomal dominant AD (ADAD, an early onset, genetic form of AD), prodromal PD, and Parkinson’s disease with dementia (PDD). Other minor categories, including carriers of specific ND-causing mutations, were also analyzed with radar plots (**Supplementary Fig. 9).** Samples from individuals with ADAD (n = 114) showed a mean AD probability of 0.65and PD mean probability of 0.23, likely capturing α-synuclein pathology known to be present in many of these samples^35^. Because of the well-understood disease progression pattern in ADAD mutation carriers, we also assessed the concordance between our model’s AD probabilities and the estimated time to AD onset, known as the EYO^16^, for presymptomatic mutation carriers. Presymptomatic carriers with fewer than ten years EYO (within ten years of expected onset based on their mutation) had significantly higher AD probabilities than those between 10 and 20 years from symptom onset (20% vs 3.6%, Wilcoxon P=2.1×10^−4^, **Supplementary Fig. 10**), suggesting that even in preclinical situations, our model is capable of differentiating those closer to disease onset based on their CSF proteomic profiles. We also observed strong performance in the prodromal PD group, within a mean PD probability of 0.94 and 96.4%. Finally, we analyzed 13 samples diagnosed as PDD, that showed a mixed PD (mean probability 0.64) and AD (mean probability 0.28) profile, supporting the presence of multiple pathologies in these individuals.

To further validate the robustness of our findings, we investigated the relationship between the CSF predictions and neuropathological phenotypes as well as cognitive assessments. Specifically, we examined correlations between predicted disease probabilities and Clinical Dementia Rating (CDR) scores, Braak neurofibrillary tangle stages, and neuritic plaque burden^36^ (C Score; **Fig. 4B**). We first analyzed AD & CO probabilities across ADAD subgroups, including affected carriers, presymptomatic carriers, and unaffected individuals, stratified by global CDR scores (CDR = 0, 0.5, and ≥1). AD probabilities increased with higher CDR scores in the DIAN cohort, with low probability of AD in individuals with CDR = 0 (mean AD probability = 5.4%), moderate for CDR = 0.5 (mean AD probability = 50%), and high for CDR ≥ 1 (mean AD probability = 81.8%, Kruskal-Wallis P = 1.99×10^−33^). A similar increase in AD probability with worsening global CDR was observed in the Knight ADRC cohort **(Fig. 4B**, Kruskal-Wallis P = 3.19×10^−198^). Consistent trends were also seen across neuropathological AD measures, Braak staging and CERAD scores, reinforcing the biological plausibility of the model’s predictions. Samples from individuals in earlier Braak stages (stages 1 & 2) had high CO probabilities (mean probability = 85.9%), consistent with isolated and lowly-abundant tau deposition in the brain. Moderate Braak stages (3 & 4) likewise showed moderate probabilities of both CO and AD, with stage 3 having slightly higher CO probability (57.3% vs 37.9% for AD) and stage 4 having slightly higher AD probability consistent with wider tau spread. In the later stages (5 & 6), almost all samples have high AD probability, consistent with widespread and abundant tau accumulation present in AD (mean AD probability = 83.9%, **Fig. 4B, panel 3**). The same trend was observed for C score, a measurement of neuritic plaque burden in the brain. In stages 0 and 1, consistent with sparse plaque presence, CO probability was high (mean CO probability in C0 = 77.7%, in C1 = 59.5%). As with Braak, the later stages of the C-score showed much higher AD probability (mean AD probability in C2 = 78.7%, in C3 = 84.7%) and lower CO probability, consistent with strong evidence of AD pathology (Kruskal-Wallis P = 1.55×10^−19^; **Fig. 4B, panel 4; Supplementary Fig. 10**). As expected, we also found low probabilities of FTD & DLB for these neuropathological scores.

**Figure 4.**
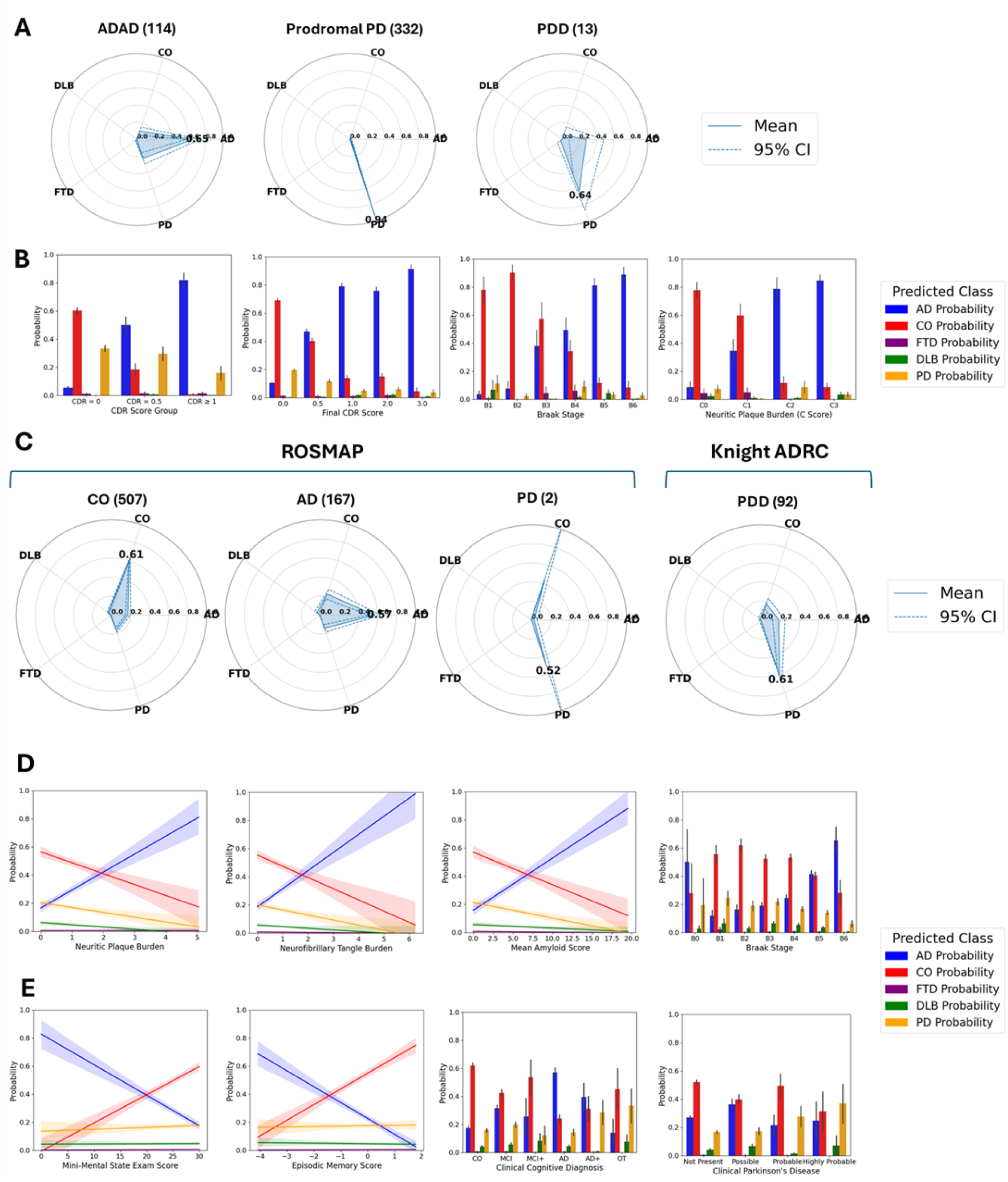
External validation and clinical pathological correlates of the proteomic classifier. (A) CSF zero-shot validation performance. Radar plots depict mean class probability profiles (AD, PD, DLB, FTD, CO) for internal CSF samples not used in training or testing: autosomal dominant AD (ADAD, n = 114), prodromal PD (n = 332) and Parkinson’s disease dementia (PDD, n = 13). **(B) CSF clinical and pathological correlations.** Predicted AD probability stratified by: Clinical Dementia Rating (CDR) at baseline within the DIAN cohort, final CDR within the internal dataset, Braak neurofibrillary tangle (NFT) stage and neuritic plaque C score. Bars show mean ± s.e.m; higher clinical or pathological burden aligns with higher AD probability. **(C) Plasma cohort level performance.** Radar plots for categories from ROSMAP or held-out diagnoses from internal data. ROSMAP: cognitively unimpaired controls (n = 507), neuropathology confirmed AD (n = 167) and PD (n = 2). Knight ADRC: ADAD carriers (n = 125) and PDD (n = 92). **(D) Concordance between model assignments and ROSMAP neuropathology findings.** Left panels: linear fits (95 % CI) of AD probability versus neuritic plaque burden and neurofibrillary tangle burden across brain regions (higher is worse AD pathology). Right panels: mean probabilities (± s.e.m) across Amyloid and Braak stage, which depend on plaque and tangle abundance. **(E) Concordance between model assignments and ROSMAP clinical findings.** Scatter plots with regression lines relate AD probability to Mini Mental State Examination (MMSE) and episodic memory zscores. Bar charts show mean probabilities (± s.e.m.) across clinical cognitive diagnosis strata and clinical PD status.

In parallel, we used other disease categories from the Knight-ADRC cohort **(Supplementary Figs. 11)** as well as independent external datasets to validate our plasma model. First, we accessed clinically-defined AD & CO samples from the Religious Orders Study – Memory and Aging Project (ROSMAP, N_CO_ = 507, N_AD_ = 167, **Fig. 4C, Supplementary Fig. 12**) and biomarker-confirmed samples from the Indiana ADRC (I-ADRC, N_CO_ = 58, N_AD_ = 32, **Supplementary Fig. 13**). In both cohorts, performance was strong for the two main categories (AD & CO). In ROSMAP, the model achieved an AUC-ROC of 0.80 for distinguishing neuropathology-confirmed AD from controls (**Fig. 4C; Supplementary Fig. 12**). Some controls (16.6%) were classified as AD, which may be due to the existence of presymptomatic molecular changes that the model is able to capture. Indeed, half of the controls that were classified as AD ended up being diagnosed with AD before death, compared to only 32% of controls classified as CO. Samples classified as CO also took approximately 2.4 years longer to develop AD than those classified as AD (mean time from blood draw to AD diagnosis = 7.3 years for those classified as CO vs 4.9 years for AD-classified samples, p=9.82×10^−5^). Similar results were seen in the I-ADRC samples. We observed very low probability of AD for these samples (mean AD probability = 9.0%), consistent with their negativity for both AD biomarkers. Interestingly, the PD probability for these samples was 0.39, suggesting that some of these clinical controls may present some PD-related pathology (**Supplementary Fig. 13**). In the Knight-ADRC, mixed pathology was also captured in individuals with Parkinson disease with dementia (PDD) as this group showing a mean PD probability = 0.61 (64.1% of PDD cases had PD as the main predicted category; **Fig. 4C, right**) and a mean AD probability of 0.18 (17.4% of PDD cases had AD as the main predicted category).

Next, because detailed neuropathological data was available for ROSMAP, we compared our prediction to these phenotypes and scores to ensure the model is capturing relevant biology. The AD probabilities derived from our model were able to capture AD pathology, as they were significantly correlated with neuritic plaques and neurofibrillary tangles (Pearson *R*>0.29; p<2.24×10^−20^, **Fig. 4D, panels 1&2**). The average plaque and tau burden was also higher in those classified as AD compared to those classified as CO (Welch’s t-test p<1.17×10^−8^,**Supplementary Fig. 14A**). Corresponding to these findings, AD probability also increased consistently with amyloid score (Pearson *R*=0.38, p=8.76×10^−25^) and was significantly higher in Braak stage 6 vs stage 1 (Wilcoxon p=1.4×10^−8^, **Fig. 4D, panels 3&4**). AD probability was also highest in samples from the “definite” CERAD score category, another measure of neuritic plaques (Kruskal-Wallis p=9.45×10^−20^; **Supplementary Fig. 10**), while for NIA-Reagan AD diagnosis, the highest likelihood AD group also had the highest model-assigned AD probability, with corresponding decreases with reduced certainty in AD presence (Kruskal-Wallis p=6.97×10^−19^, **Supplementary Fig. 15**). Participants classified as AD by the model also exhibited significantly higher Braak neurofibrillary tangle stages compared to those classified as CO, with a mean Braak stage of 4.25 in the predicted-AD group versus 3.73 in the predicted-CO group (Welch’s t-test p=8.99× 10⁻⁹; **Supplementary Fig. 14C**). These trends generally held for other neuropathological and disease-related phenotypes as well (**Supplementary Fig. 10A**).

We next sought to extend the analyses of our predicted categories and probabilities to cognitive and clinical examination phenotypes in the ROSMAP cohort. Our AD probabilities showed a significant negative correlation with Mini-Mental State Exam (MMSE) scores (Pearson *R*=−0.38; p=2.18×10^−35^) and with episodic memory performance (Pearson *R*=−0.37; p=8.96×10^−31^). On the other hand, CO probabilities showed the opposite pattern to that of AD-probabilities (Pearson *R*>0.32; p<1.21×10^−23^). No association was found between these cognitive scores and the PD, FTD or DLB-probabilities (**Fig. 4E, panels 1&2**, absolute Pearson *R*<0.04, p>0.22). Other cognitive tests, including other forms of memory and cognitive function, showed similar patterns (**Supplementary Fig 15)**. Similarly, AD probability increased systematically along the clinical continuum from no impairment through MCI to AD (**Fig. 4E, panel 3,** MCI vs NCI Wilcoxon p=1.52×10^−10^, AD vs MCI p=3.90×10^−11^, Kruskal-Wallis p=1.74×10^−32^). While these results support our model’s ability to predict AD, the ROSMAP dataset also included individuals with evidence of PD. We also observed good concordance between these PD phenotypes and corresponding PD probability. Those with highly probable PD based on clinical assessment had the highest PD probability, while other categories (no evidence, possible, and probable PD) had lesser PD probabilities consistent with their level of confidence (**Fig. 4E, panel 4;** Kruskal-Wallis p=8.78×10^−3^). Analyses of PD-relevant clinical examinations also confirmed the ability of the model to capture PD pathology, as samples classified as PD had higher Global Parkinson’s Summary scores^37^ than those classified as controls (13.29 in PD-classified samples, 9.84 in CO-classified samples; p=5.42×10^−3^; **Supplementary Fig. 14D).**

Taken together, these data demonstrate that our proteomic models can accurately classify individuals in the most likely category without training or imputation and that the assigned probabilities align quantitatively with gold-standard neuropathological and cognitive measures, underscoring their potential utility for research and clinical stratification.

#### AI-based Reclassification of Ambiguous Clinical Diagnosis

Based on our validation of the main disease categories and zero-shot performance using neuropathological and cognitive test data, we next sought to determine if we could use our model to reclassify individuals with unclear diagnosis into more appropriate diagnostic groups. This application of AI-based models holds significant promise, because they may be able to capture changes in individuals at risk of disease development well before symptoms actually develop^16^.

Because we used a strict definition for control samples in the training of our CSF model (based on amyloid & tau negativity), we first attempted to identify samples from the Knight-ADRC who may not have any clinical symptoms but may have underlying AD pathology. In total, the Knight-ADRC included 1,534 CSF samples obtained from individuals who were clinically defined as controls. We first analyzed biomarker information for this group. Using a six-month time window around the CSF draw date, 912 of the samples had corresponding AD CSF biomarker information (amyloid & tau). The majority of these were, as expected, negative for both biomarkers (A-T-, N=558), but a mix of other profiles were also present (N_A+T-_=207, N_A-T+_=55, N_A+T+_=92), suggesting a substantial proportion of these samples may have underlying disease pathology that our model may be able to capture. We first compared the classification signatures identified by the AI model between the biomarker groups. The A-T-group was almost exclusively classified as controls (550/558, 98.6%; mean AD probability = 0.5%, **Fig. 5A, radar plot**), with only one sample classified as AD, providing evidence of very high concordance between the model and the gold standard AD biomarkers. The two intermediate groups (A+T-, A-T+) both had comparatively higher proportions of AD-predicted samples (A+T-: 14.0% classified as AD, 75.8% as CO, mean AD probability = 13.4%; A-T+: 50.9% AD, 49.1% CO, mean AD probability = 51.6%). This trend continued with the individuals who were positive for both amyloid and tau (A+T+), with 78.3% of these samples classified as AD while only 18.5% were assigned as CO. We further analyzed the 18.5% cognitive normal individuals that were A+T+ but were still classified as CO by our classifier. Of the 17 samples in the A+T+, CO-classified group, 11 were still diagnosed as controls upon their last follow-up (maximum of 7.83 years after draw date), supporting their classification as controls. The other five eventually developed AD; four became cases at least five years after the CSF drawdate, while one was diagnosed two years after. The substantial time gap between CSF drawdate and diagnosis for 4 of 5 of these individuals suggests their classification as controls is supported by clinical evidence.

**Figure 5.**
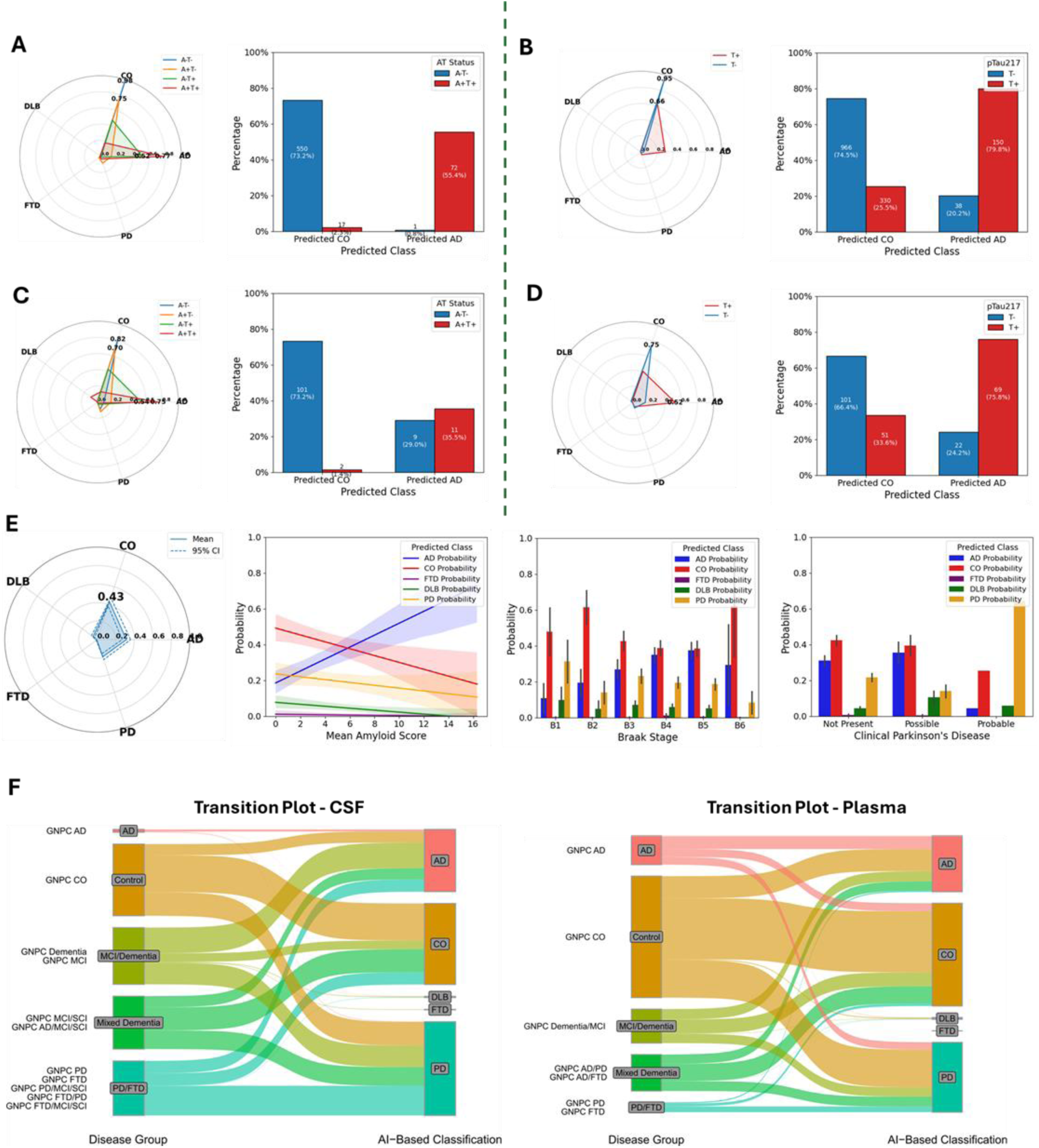
AI-based reclassification of ambiguous clinical diagnosis. (A) CSF clinical controls. *Left:* Radar plot showing mean class-probability profiles (AD, PD, DLB, FTD, CO) for participants clinically labeled as CO at enrollment, separated by CSF AD biomarker status (amyloid and tau, AT). *Right:* Bar charts display the distribution of CSF AT status among samples the model retained as CO versus those reclassified as AD. **(B) Plasma clinical controls.** Radar plot and bar charts analogous to panel A, using plasma-based class probabilities and pTau217 positivity (T+/T–). **(C) CSF “Other” diagnoses (OT).** Same analyses as in panel A, applied to individuals with indeterminate clinical labels. The radar plot reveals mixed probability profiles, while bar charts compare AT status between samples reclassified as CO versus AD. **(D) Plasma “Other” diagnoses.** Radar plot and bar charts stratified by pTau217 status illustrate concordance between AI-based classification and biological markers. **(E) ROSMAP mild cognitive impairment (MCI).** Panel 1: Radar plot of class probabilities for MCI participants. Panels 2 & 3: AD probability plotted against amyloid PET Centiloid scores and post-mortem Braak NFT stages. Panel 4: Mean class probabilities (± s.e.m.) stratified by clinical Parkinson’s disease certainty levels. **(F) Transition plots.** Samples from the GNPC are shown with their clinical diagnoses (left) and their AI-based classifications (right) in CSF and plasma.

We next looked at the inverse relationship, focusing first on AI classification and then diving into the biomarker profiles within the AD and CO classification groups. Those classified as CO were predominantly negative for CSF biomarkers (73.2% A-T-, **Fig. 5A, panel 2**) and for brain imaging-based amyloid detection (85.3% amyloid PET-negative, **Supplementary Fig. 16**). The opposite trend was observed for the AD-classified samples. In that group, 71.0% were positive for amyloid PET (**Supplementary Fig. 16**) and 55.4% were A+T+ **(Fig. 5A, panel 2).** More importantly, of the 130 AD-classified, control-diagnosed samples with CSF biomarker information, only one was A-T-, suggesting our model is able to highly accurately capture AD-relevant molecular changes before symptom development.

We next focused on the reclassification of the plasma samples obtained from clinically defined controls. While CSF AD biomarker or amyloid-PET information was unavailable for most of these individuals, plasma pTau217 was available for a majority of the samples and more robust biomarker profiles were available for a smaller subset. We again observed good concordance between our classifications and biomarker status^38^ (**Fig. 5B, bar plot**). Clinically defined controls that our model classified as CO were mainly pTau217-negative (966/1296, 74.5%), while those classified as AD were overwhelmingly pTau217-positive (79.8%, **Fig. 5B**). Using amyloid-PET provided similar findings, with 84.4% of classified COs also having negative amyloid-PET results compared to only 22.2% of those classified as AD **(Supplementary Fig. 16).** Therefore, across imaging, blood, and CSF biomarkers, our plasma classifier showed good accuracy for classifying samples into CO and AD categories.

Some individuals may present symptoms of a neurodegenerative disease without a clear clinical diagnosis, represented in our cohorts as “other” (labeled as OT). Clarifying their diagnoses using our AI-based model may lead to improvements in their care and their ability to plan for the future. Given their unclear diagnoses, OT samples (**panel 5C & D**) showed mixed probability profiles. Across the 266 OT samples in CSF, 159 were classified as CO, 48 as AD, 6 as DLB, 17 as FTD, and 36 as PD. We first assessed the concordance between model classification and AD biomarker status. For those classified as CO (N=138), 73.2% were also negative for both AD biomarkers in CSF (A-T-, Fig. 5C), while only 1.4% of the CO classifications were A+T+. Of OT samples classified as CO with amyloid-PET, 73.3% of them were negative for amyloid on brain scans (**Supplementary Fig. 16**). Only one of the two A+T+ CO-classified samples had follow-up information; as of six years after the CSF draw, they were re-considered a clinically defined control, supporting our CO classification even with positive AD biomarkers. On the other hand, OT samples classified as AD (N=31) had higher rates of biomarker positivity (71.0% positive for at least one biomarker, **Fig. 5C**) and amyloid-PET positivity (71.4% amyloid-PET positive, **Supplementary Fig. 16**). Notably, follow-up clinical or neuropathological data was available for a subset of the samples; 43 were re-diagnosed as controls on follow-up, of which 33 were classified by our model as CO. All others were still considered OT at follow-up.

In plasma, 284 samples from the Knight-ADRC did not have a clear clinical diagnosis (OT). The majority were classified as either CO (59.1%) or AD (35.6%), with only 15 samples classified into the other three groups. We again observed good concordance with AD-relevant biomarkers. Overall, 152 CO-classified samples had pTau217 information, with 66.4% of those negative for the biomarker. Conversely, 75.8% of the 91 AD-classified samples were positive for pTau217 (**Fig. 5D**). A subset of the samples also had follow-up clinical data. While most were still diagnosed as OT, 33 samples were re-diagnosed as controls, with 25 of those also being classified by our model as CO, further supporting its accuracy. To confirm that reclassification using our model leads to high quality findings, we investigated samples from individuals diagnosed with mild cognitive impairment (MCI) from the ROSMAP cohort (**Fig. 5E).** These samples were generally classified as either CO (n=115), AD (n=86), or PD (n=45). Within this group, we first looked at measurements of AD-related pathology. We saw significant concordance between our model’s AD probability and amyloid levels across brain regions (Pearson *R =* 0.31, p=4.97×10^−6^) and corresponding negative correlations between CO probability and amyloid (Pearson *R* = −0.17, p=0.02), while the likelihoods for PD, DLB and FTD were low (**Fig. 5E**). We next analyzed Braak stage and observed consistent increases in AD probability from stage 1 to 5 consistent with increasing pathology along the spectrum (AD probability for Braak score 1 = 10.9%, for Braak score 5 = 37.5%, Kruskal-Wallis p=0.03, **Fig. 5E**). Because information for Parkinson’s disease was also available in this cohort, we investigated the concordance between our probabilities and the clinical likelihood of PD diagnosis. Samples assigned high probability of PD based on clinical assessment in fact had high PD probabilities (Prob(PD) = 64.4%, **Fig. 5E, panel 4**), indicating that the model can predict not only AD and CO but also PD.

Finally, we applied our model (again, without any retraining) to CSF and plasma samples from the Global Neurodegeneration Proteomics Consortium (GNPC), which includes datasets from a broad range of sites with varying disease focuses, diagnostic criteria, and availability of confirmatory data, representing a worst-case scenario for application of our model (**Fig. 5F**). First, focusing on CSF, we grouped the GNPC samples from five contributors into five groups (AD, CO, unclear dementia, potential mixed dementia, and PD/FTD, **Supplementary Tables 1B & 12**). Our classifier showed AUCs of 0.87, 0.70, and 0.65 for AD, CO, and PD, respectively (**Supplementary Figs. 17, Supplementary Tables 5 & 6**). In this dataset, many samples are classified as “dementia”, denoting individuals with unclear underlying diagnosis. Of these, 186 (45.3%) samples were classified as AD, 156 (38.0%) were classified as PD and 62 (15.1%) as CO (**Supplementary Fig. 17**). As there was not additional longitudinal or clinical data for these samples, we were not able to fully validate these predictions. Additionally, a number of samples with a clinical diagnosis of CO were reclassified as AD (15.5%). Based on the earlier analyses presented here, it is likely these are presymptomatic individuals. Notably, as with the other cohorts, we observed strong concordance between predicted probabilities and AD-relevant phenotypes. First, in individuals with global CDR measurements, those with CDR≥1 had significantly higher AD and lower CO probabilities than those with CDR=0 (p<1.31×10^−35^, **Supplementary Fig. 18).** We observed similar results for MMSE, where an inverse correlation was observed between MMSE score and AD probability (Pearson *R*=−0.31, p=8.38×10^−31^, **Supplementary Fig. 18).**

In the GNPC plasma analysis, we observed strong performance, albeit not to the same degree as ROSMAP or the I-ADRC **(Fig. 5F),** likely as the GNPC is based only clinical diagnosis and do not have biomarker or neuropath data. AUCs ranged from 0.63 for FTD to 0.74 for PD, consistent with the findings from the internal testing dataset (**Supplementary Fig. 19).** While many AD samples were classified as AD (45.8%), we observed a relatively high frequency of reclassification as CO (27.5%) or PD (25.7%), suggesting some samples may be misdiagnosed. Nonetheless, we again observed strong concordance between global CDR and AD/CO probabilities, with AD probabilities significantly higher in samples from individuals with CDR ≥ 1 (p=1.43×10^−54^, **Supplementary Fig. 18**) and showing an inverse correlation with cognition as measured by MMSE (Pearson *R*=−0.21, p=1.62×10^−44^, **Supplementary Fig. 18).**

Overall, we showed across both internal and external data that our classifier has the ability to reassign diagnoses that correspond accurately with AD biomarker, neuropathology, and cognitive testing information, demonstrating the appeal of AI-based approaches to improving disease diagnosis. While performance in the worst-case scenario (GNPC) was moderate, with further model advances, these techniques may represent valuable tools for use in the clinic. As evidence of this fact, we next highlight a few specific examples of the appeal of these types of models.

### AI-based Classification on an individual basis: a step towards precision medicine

While we have shown how the model performs across large cohorts on a broad scale, it is important to determine the clinical impact at the individual level. Therefore, we selected representative examples from samples with extensive postmortem documentation available in the cohort to demonstrate that our model provides critical evidence that increased accuracy over clinical diagnosis.

First, we identified three clinically diagnosed controls from ROSMAP with AD probability greater than 0.99 according to the model, representing high confidence in AD likelihood. Each of these individuals developed AD within four years of the blood draw, suggesting molecular changes leading to AD were being accurately captured by the model. Each of them also presented at autopsy with at least Braak stage 5, representing extensive tau pathology consistent with AD.

They also had CERAD scores of 1, consistent with definite presence of neuritic plaques consistent with amyloid pathology, and were classified as high-likelihood of AD based on the NIA-Reagan diagnosis criteria. Each also had MMSE scores consistent with dementia (MMSE < 24) at their last cognitive assessment. We also identified three individuals (two diagnosed at plasma draw as AD, one as CO) that the model assigned with high confidence as DLB, contrary to the diagnosis for each individual. For all three of them, postmortem neuropathological examination confirmed a re-diagnosis of DLB based on the presence of alpha-synuclein deposits in the neocortical brain region. Two of these individuals were also assigned to Braak stage 1 and were given a low likelihood of AD according to the NIA-Reagan scale, consistent with pure Lewy body pathology. Furthermore, two samples from ROSMAP with an unclear clinical diagnosis had high AI-based probability of PD (0.99 and 0.89). Both individuals showed at least possible PD diagnosis based on clinical assessment, with high global PD summary scores^37^ consistent with the motor symptoms involved in PD. These DLB and PD examples suggest that the model uncovers latent synucleinopathy not detected by initial clinical assessment. Finally, we identified two individuals in the “OT” group with likely diagnoses of PD (**Fig. 4C**). One individual, assigned by our model to PD with a probability of 98%, was considered to have a high probability of PD, with substantial motor deficits and a moderate global Parkinson’s score supporting their diagnosis. The other individual, with a model-assigned PD probability of 4.7% (CO probability = 89.7%), was considered to have probable PD at the clinical assessment associated with the plasma draw and the one after, but at the final clinical visit the individual was not considered to have PD. At all other visits, a PD diagnosis was considered possible at best and more frequently PD was considered not present, generally supporting the model’s assignment of the individual as a CO.

Across CSF and plasma, proteomic probabilities proved capable of realigning ambiguous or transitional diagnoses and are supported by underlying biomarker status, neuropathology and cognitive assessment. These data emphasize the classifier’s potential to refine clinical categorization, highlight mixed pathologies, and guide appropriate downstream management even when conventional diagnostic labels are inconclusive or inaccurate.

## Conclusion / Discussions

As the world population continues to age and the prevalence of neurodegenerative diseases increases^36^, the identification of biomarkers for these debilitating conditions has been a highly active area of research. Established success to this point has only been obtained in the field of Alzheimer’s disease, with the implementation over the last decade of the amyloid/tau/neurodegeneration paradigm and the recent development of the first plasma-based AD biomarker using pTau217^29,30,39^. Most biomarker identification studies focus solely on a single form of dementia, limiting the broad relevance of these approaches^34^. Specifically, most proteomic studies for Alzheimer’s disease have been focused on CSF and have identified novel biomarkers such as SMOC1 or 14-3-3 protein that show good predictive power for AD^9,10,16,40,41^, while recent studies have identified DDC as a strong candidate CSF biomarker for PD^42,43^ and some candidate biomarkers for FTD^44^. While blood biomarkers are preferable to those in CSF due to the less invasive sample collection, recent studies mainly focus on one disease^34,45^ which makes it difficult to determine if the identified biomarkers are specific to one type of dementia or if they reflect common neurodegenerative processes. Although there are some studies that have included more than one type of neurodegenerative disease, they are limited in sample size and the cross-disease comparison is normally limited in scope^34,46^.

Given the frequent symptomatic and pathological overlap across these diseases^47,48^, identifying biomarkers and models that can discriminate between different ND types is crucial. With the advent of artificial intelligence approaches that allow for the concurrent analysis of not only multiple data types but also of multiple disease types, biomarker discovery is primed to expand. Here we designed, tested, and validated a highly accurate multi-disease classifier model across multiple independent cohorts solely based on proteomic data from CSF or plasma. Our multi-class framework can capture overlapping protein signatures across multiple neurodegenerative categories by providing multi-status joint probabilities, which also allow for interpretation as to the confidence of the classification. Our use of a single widely available data type proteomics substantially simplifies the process involved in model development and reduces cost to the patient. Our explainable model also allows for investigation of the individual proteins that contribute to the assigned outcomes. As evidence of this interpretability, we identified established AD biomarkers (YWHAG, NPTX2) and known treatment targets (ACHE) as strongly associated with AD/CO probability (**Supplementary Figs. 6 & 7**)^9,32,33^, suggesting our model is capturing known biological signals and adding additional information.

We validated the performance of our model across cohorts, observing AUCs above 0.95 in our CSF testing set and close to 0.90 in the plasma testing set for AD, PD, and CO classes, with model performance comparable to that of pTau217, the gold-standard AD biomarker in plasma^39^, and amyloid PET, a key imaging test for AD. Overall, we observed macro AUCs of 0.95 in CSF and 0.79 in plasma, supporting the accurate classification of samples. Performance continued to be strong in external cohorts, where we observed AUCs around 0.80 in ROSMAP and 0.7-0.8 for AD, PD, and CO in the GNPC, highlighting the cross-cohort applicability of our CSF & plasma models. Notably, we are able to obtain this level of accuracy while also differentiating between multiple types of ND, making the model accuracy all the more relevant compared to single-disease biomarkers like pTau217 or CSF amyloid/tau status.

Using neuropathological, imaging, and biomarker data, we demonstrated strong concordance between our model and the biology underlying disease. Our model was able to accurately classify samples from individuals with ADAD, prodromal PD, or PDD, representing a difficult task for AI-based classifiers because no samples from those populations had previously been seen by the model to form a reference point. This suggests that it may be able to group individuals with rare or emerging forms of ND into their most-similar diseases to assist with treatment and patient counselling.

A key finding from our analysis was the ability of the model to reclassify individuals with underlying disease hallmarks or unclear etiology (**Fig. 5**). While biomarkers are making this more possible, there is still a serious need for methods that can identify those at risk of dementia development. AI-based models could be valuable for participant recruitment in clinical trials and for assisting clinicians in providing complete information relevant to life planning for those at risk of dementia development. Additionally, our datasets highlight that a substantial number of individuals with cognitive decline have unclear clinical diagnoses, potentially limiting the therapeutic options available to them. Our model showed the ability to determine the most likely form of dementia in many of these individuals, which may improve their care options and increase their quality of life.

Our model exhibits a number of benefits over previous approaches that improve its scalability and wide applicability. First, our model’s use solely of proteomic data is a much simpler and less expensive approach than many other AI-based models that require integration of several expensive datatypes such as neuroimaging data^50–53^, electrical medical records, medication history, and clinical data^23,53,55^. The use of fluid proteomics, on the other hand, is not only more affordable but also depends solely on a blood or CSF draw. While CSF draws are relatively invasive, blood draws are quite trivial, can be done at satellite clinics, and do not require extensive training or expertise to perform, making plasma a vastly more accessible resource than neuroimaging data. Additionally, unlike our multi-disease model, many previous approaches focus on a single disease^50,56^, which may limit their ability to capture co-pathology or common disease processes across NDs^57^. There are a few studies that have performed multi-disease analyses, but these frequently classified samples simply using a binary-focused paradigm (disease yes/no)^58^, as opposed to the multi-class probability-based approach of our model that substantially increases the granularity of our output. The continuous nature of our estimates provides substantially more information than a binary classification, as shown by the trend of increasing AD probability with more AD-like biomarker profiles (**Fig. 5A**) and with proximity to the expected age of AD onset in presymptomatic carriers of autosomal dominant AD mutations (**Supplementary Fig. 10**). While one study has similarly attempted to develop a model that assigns multi-disease probabilities through an AI approach^23^, the model was built through the incorporation of imaging, demographics, medical history, medications, and cognitive assessments, substantially increasing the complexity compared to our simple proteomics-based model. Even with these diverse forms of data, our model demonstrated equivalent performance (multimodal model AUC = 0.94 vs approximately 0.9 using proteomics for similar outcomes), demonstrating the robust nature of proteomic data. Because of our use of an explainable tree-based gradient boosting model, our approach also has benefits over “black-box” approaches such as neural networks (NNs) in interpretability^59^. In contrast, NN-based approaches often lack transparency in layer-wise transformations and do not provide sufficient explanations for how features contribute to the final prediction, as the contribution of each original feature to the end products is largely lost because of the intermediate layers. Gradient boosting models have the benefit of metrics such as feature importance or SHAP values^60^, which allow for the interpretation of the importance of each feature to the overall model. Similar metrics are not available for neural networks or transformer models. Additionally, by building our model in a single step through the integration of all samples, we reduce biases that are introduced when multiple models (i.e., AD vs CO, PD vs CO, FTD vs CO) are combined post-hoc, increasing the robustness and confidence of our model. Our model also showed a robust ability to be applied to external datasets without the need for retraining, which greatly simplifies the ease in which it can be implemented across cohorts.

Although our approach has major strengths over previous efforts, we acknowledge that weaknesses remain. While performance for the more common classes (AD, PD, and CO) is strong and robust across cohorts, the small sample sizes used for FTD and DLB and the known overlap between them and other forms of neurodegenerative disease limit the accuracy with which they can be classified due to a lack of proteins that are robustly and consistently associated with these diseases. However, as a first effort towards this approach, we have shown the appeal of the technique and emphasize that expansions of sample sizes for these less common forms of dementia should greatly improve model accuracy. Importantly, validation of our model using biological evidence was largely limited to AD and was limited to certain cohorts, because relevant pathology and imaging data was not available for the other disease types or in many samples. This could impact the initial accuracy of our model and limits our ability to assess the accuracy of its performance in outside cohorts that lack robust underlying information. In the future, post-mortem analysis of individuals with FTD & DLB diagnoses should be used to increase confidence in model performance. Additionally, one of the main problems of proteomic-based approaches is the normalization of data across cohorts, batches, and sample acquisition procedures, that can lead to biases in the prediction^9,12,61^. Improved normalization procedures will be needed to ensure biological context is kept while cross-cohort differences are minimized. Finally, the weaker performance in the GNPC compared to other cohorts could be due to a combination of cohort composition differences and inaccuracies in clinical diagnosis, which would lead to an inaccurate baseline diagnosis to compare to our classification.

In summary, we demonstrated the ability of our models to reclassify individuals with underlying pathological disease hallmarks into disease states that may clarify their medical care. Importantly, we showed that disease probabilities tracked accurately with gold-standard biology from neuroimaging and cognitive tests, lending credence to the model. The model was able to properly interpret and classify unseen disease types (Zero-shot), such as such as autosomal dominant AD (ADAD) and Parkinson’s disease with dementia (PDD), without requiring additional labeled data or imputation. We further highlighted specific examples where our classifier accurately assigned individuals to a specific status based on underlying pathological data that differed from their clinical diagnosis. Collectively, these efforts enhance both clinical decision-making and research into the complexity of dementia, offering a benchmark computational framework that captures the multifaceted nature of neurodegenerative disorders.

## Methods

### Cohort Overview

To maximize participant diversity and ensure rigorous external validation, we assembled a cross-sectional dataset from both local and external neurodegenerative cohorts, drawing on each individual’s last available sample collection date (**Table 1**). The local cerebrospinal fluid (CSF) dataset comprised 4,677 samples from the Charles F. and Joanne Knight Alzheimer’s Disease Research Center^62^ (Knight ADRC, n=1,055), Stanford ADRC (n=264), Movement Disorder Clinic at Washington University (MDC, n=238), Barcelona-1 (n=206), ACE (n=620), PPMI (n=1,075), DIAN (n=482), and ADNI (n=737). These cohorts were measured on the SomaScan 5k (PPMI, Stanford) or 7k platforms (all others), prompting subsequent harmonization of overlapping analytes. In parallel, 4,750 cross-sectional plasma samples were derived from the Knight ADRC (n=3,110), Stanford ADRC (n=628), and MDC (n=1,012), all on the SomaScan 7k platform.

The Charles F. and Joanne Knight-ADRC at Washington University School of Medicine (St. Louis, MO. USA) recruits and longitudinally assesses community-dwelling adults older than 45 years via prospective studies of memory and aging since 1979. All studies were approved by the Human Research Protection Office at Washington University, and written informed consent was obtained from all participants. The Memory and Aging Project at the Knight ADRC (Knight ADRC-MAP) involves longitudinal collection of biofluids (plasma, CSF, fibroblast), annual clinical assessments, neuropsychological testing, and neuroimaging studies, as well as collection of autopsied brain samples. Eligible participants may be asymptomatic or have mild dementia at the time of enrollment. All participants are required to participate in core study procedures, including annual longitudinal clinical assessments, neuropsychological testing, neuroimaging, and biofluid biomarker studies. Annual assessments of the participants were performed by experienced clinicians using a semi-structured interview with knowledgeable collateral source and the symptomatic individual in accordance with the Uniform Data Set protocol of the National Alzheimer’s Coordinating Center^63^, as well as a detailed neurological examination. Participants comprise Non-Hispanic White individuals from North America (82.5%) and African-Americans (13.3%). Samples have been obtained from over 5,510 participants, including 2,426 AD cases, 148 FTD, 88 DLB, and 2,156 cognitively normal healthy individuals. Autopsy material are available for over 1,182 participants including 474 with fresh frozen parietal brain tissue (https://dss.niagads.org/datasets/ng00127/). Multi-tissue (brain, CSF, and plasma), multi-omics data (genetics, epigenomics, transcriptomics, proteomics, and metabolomics) have been generated for the purpose of identifying novel risk and protective variants for dementia and potential drug targets. Participants from the Knight ADRC were included in this study if they were cognitively unimpaired with a global clinical dementia rating (CDR) score of 0 at enrollment. A clinical diagnosis of incident dementia is considered by study clinicians at the conclusion of each annual assessment, integrating results from the clinical assessment and bedside measures of cognitive function^64^. Dementia was diagnosed according to the National Institute of Neurological Disorders and Stroke criteria^65^ and National Institute on Aging-Alzheimer’s Association Work Group criteria for participants assessed after 2011^66^. Diagnosis of AD dementia was made in accordance with criteria developed by working groups from the National Institute of Aging and the Alzheimer’s Association^66^. Diagnosis of vascular dementia conformed to the NINDS-AIREN criteria^67^. More information is available at knightadrc.wustl.edu.

Samples were acquired through the National Institute on Aging (NIA)-funded Stanford Alzheimer’s Disease Research Center (SADRC). The SADRC cohort is a longitudinal observational study of clinical dementia subjects and age-sex-matched non-demented subjects. The collection of plasma was approved by the Institutional Review Board of Stanford University and written consent was obtained from all subjects. Blood collection and processing were done according to a rigorous standardized protocol to minimize variation associated with blood draw and blood processing. Briefly, about 10 cc whole blood was collected in a vacutainer EDTA tube (BD Vacutainer EDTA tube) and spun at 3000RPM for 10 mins to separate out plasma, leaving 1 cm of plasma above the buffy coat and taking care not to disturb the buffy coat to circumvent cell contamination. Plasma processing times averaged approximately one hour from the time of the blood draw to the time of freezing and storage. All blood draws were done in the morning to minimize the impact of circadian rhythm on protein concentrations. All healthy control participants were deemed cognitively unimpaired during a clinical consensus conference that included board-certified neurologists and neuropsychologists. Cognitively impaired subjects underwent Clinical Dementia Rating and standardized neurological and neuropsychological 5 assessments to determine cognitive and diagnostic status, including procedures of the National Alzheimer’s Coordinating Center (https://naccdata.org/). All participants included in this study were deemed cognitively impaired during a clinical consensus conference that included neurologists and neuropsychologists. All participants were free from acute infectious diseases and in good physical condition. Samples were also obtained from the Stanford Aging Memory Study (SAMS). SAMS is an ongoing longitudinal study of healthy aging. Blood collection and processing were done by the same team and using the same protocol as in SADRC. Neurological and neuropsychological assessment were done by the same team and using the same protocol as in SADRC. 192 participants were included in the present study, and 11 were participants in both the SADRC and SAMS study.

The Movement Disorder Clinic at Washington University recruits participants with Parkinson’s Disease (PD) and other movement disorders, from whom DNA, blood RNA, plasma, and CSF is collected. The goal of the study is to identify early disease biomarkers as well as markers for memory decline in the PD population^68,69^.

Cerebrospinal fluid samples were obtained from Barcelona-1, a study led by the University Hospital Mutua de Terrassa in Terrassa, Spain. Barcelona-1 is a longitudinal study consisting of about 300 individuals, who underwent PET and cerebrospinal fluid collection if determined to have mild cognitive impairment (MCI) or greater. Individuals underwent follow-up analyses to track the progression of disease. People included in the study consist of those diagnosed with AD dementia, non-AD dementias, MCI, or subjective memory complaints.

We further obtained cerebrospinal fluid samples from Ace Alzheimer Center Barcelona (ACE), a private non-profit group dedicated to the diagnosis, treatment, and assistance of patients and families, and to the study of Alzheimer’s disease. Headquartered in Barcelona, Ace was founded in 1995 and has diagnosed over 30,000 patients, collected 20,000 blood, 9,000 plasma, and 3,000 cerebrospinal fluid samples, analyzed almost 13,000 genetic samples^70,71^, and participated in over 150 clinical trials so far. For more details, visit www.fundacioace.com/en.

The Parkinson’s Progression Markers Initiative (PPMI) is a large study focused on the investigation of biological markers of Parkinson’s Disease (PD). Founded in 2010 with support from the Michael J. Fox Foundation, PPMI plans to enroll about 4,000 people in intensive clinical and imaging testing and about 50,000 more for genotyping and simpler PD-related tests. Individuals undergo cerebrospinal fluid draw, MRI, motor assessments, and many more tests to get a comprehensive view of PD-related phenotypes. For more details on PPMI, please visit www.ppmi-info.org.

Cerebrospinal fluid and plasma samples were obtained from the Dominantly Inherited Alzheimer’s Network (DIAN). Led by the Washington University in St. Louis School of Medicine, DIAN utilizes a family-based long-term cohort study to investigate Autosomal Dominant Alzheimer’s Disease (ADAD). Tissues collected (blood, CSF) are analyzed to detect changes in carriers of mutations causal for ADAD. The samples and data utilized in this study are from the 15^th^ datafreeze (DF15). For more information about DIAN, visit dian.wustl.edu.

Cerebrospinal fluid samples used in the preparation of this manuscript were obtained from the Alzheiomer’s Disease Neuroimaging Initiative (ADNI) database (<u>adni.loni.usc.edu</u>). ADNI was launched in 2003 as a public-private partnership, led by Principal Investigator Michael W. Weiner, MD. The primary goal of ADNI has been to test whether serial magnetic resonance imaging (MRI), positron emission tomography (PET), other biological markers, and clinical and neuropsychological assessment can be combined to measure the progression of mild cognitive impairment (MCI) and early Alzheimer’s Disease (AD).

For independent external validation, we leveraged the Global Neurodegeneration Proteomics Consortium (GNPC) v1.3, a large-scale, multi-institution initiative comprising more than 40,000 CSF and plasma samples collected worldwide. Specifically, GNPC provided 3,050 cross-sectional CSF samples from five contributors (“N,” “Q,” “T,” “O,” “J”), where N, Q, and T used the CSF_7k panel, and O and J used CSF_5k. In addition, GNPC included 17,901 cross-sectional plasma samples, of which 6,103 EDTA_Plasma_7k samples (contributors “Q,” “G,” “A,” “I,” “J,” “L,” and “R”) were selected to ensure broad representation of neurodegenerative statuses and sufficient sample sizes. We excluded contributor F and a subset of J from GNPC to avoid duplicating participants already represented in the Knight-ADRC or Stanford ADRC (**Supplementary Table 12**). GNPC encompasses an extensive range of clinical diagnoses and staging levels, including cognitively healthy controls (CO), AD, PD, FTD, amyotrophic lateral sclerosis, and other syndromes. Where detailed clinical diagnoses were unavailable, GNPC data harmonization relied on Clinical Dementia Rating (CDR) and Mini-Mental State Examination (MMSE) scores to categorize individuals as control, mild cognitive impairment (MCI), or dementia. For participants with available CDR scores, we applied the following criteria: CDR = 0 was classified as ‘CO’; CDR = 0.5 as ‘MCI’; and CDR > 0.5 as ‘Dementia’.

We also analyzed samples from ROSMAP. The Religious Orders Study^72^ and the Rush Memory and Aging Project (collectively ROSMAP) are community-based, longitudinal cohorts that follow older adults who have consented to annual cognitive evaluations and brain donation at death. ROS recruits members of U.S. religious communities (est. 1994); MAP enrolls residents of retirement facilities and private homes in the Chicago area (est. 1997). Standardized post-mortem protocols quantify Alzheimer’s pathology, Lewy bodies, infarcts and other age-related lesions. For this work we analysed SomaScan 7k proteomes from 973 participants: 507 cognitively normal controls, 167 neuropathology-confirmed AD cases and 17 cases with AD plus an additional contributing pathology. All procedures were approved by the Rush University Medical Center IRB, and written informed consent plus an Anatomic Gift Act were obtained from each participant.

Finally, we included samples from the Indiana ADRC in our external validation. The Indiana Alzheimer’s Disease Research Center (IADRC) cohort includes individuals with Alzheimer’s disease (AD) dementia and cognitively normal (CN) elderly controls enrolled through the Indiana Memory and Aging Study (IMAS)^66^. All participants gave informed consent under IRB-approved protocols. They underwent standardized neurological and cognitive evaluations (UDS-3) and provided blood samples for biomarker profiling. Diagnoses followed established AD criteria, and CN participants showed no significant cognitive impairment. The cohort is racially diverse, including White, Black or African American, and Asian individuals. This study included 103 IADRC participants with available proteomic and clinical diagnosis with the ATN information: 58 COs with A-T-, 12 COs with A+T-, 1 CO with A+T+, and 32 AD with A+T+.

### Proteomic Platforms and Baseline Quality Assessments

All cohorts in this study were profiled using the SomaScan platform^73^, an aptamer-based technology recognized for its high sensitivity (femtomolar to micromolar) and reproducibility (median intra- and inter-plate coefficients of variation ∼5%). Depending on the cohort, either the 5k or 7k panel was used, capturing up to 7,584 aptamers that target thousands of human proteins. SomaLogic performed baseline normalization, including hybridization controls for intra-plate variation, median-signal corrections across plates, and additional normalization against an external reference pool to address potential batch effects. Most CSF samples were obtained by morning lumbar puncture in fasting or near-fasting conditions, while plasma samples were collected with some variations in fasting status and collection times. Each cohort adhered to standardized collection protocols, and all samples were shipped on dry ice to SomaLogic for protein quantification.

To ensure data integrity, we performed additional quality control (QC) steps at both the analyte and sample levels as detailed previously^16,74^. For plasma, we removed analytes for which 85% of the total samples failed the limit of detection (LOD) filter, defined as the mean relative fluorescent units of the analyte + 2*SD in buffer samples. Due to lower protein abundance in CSF, no LOD filter was applied. Analytes were excluded if they targeted non-human proteins or if their calibration scale factors (SF) deviated by more than 0.5 across plates. Analytes exceeding a median coefficient of variation (CV) of 0.15 were also removed. Individual analyte measurements with values more extreme than Q3+1.5*IQR or Q1-1.5*IQR, where Q1/Q3 is the 1^st^ and 3^rd^ quantile value for that analyte across samples and IQR is the interquartile range, were considered outliers and were replaced with NA values. Analyte call rate was calculated and analytes with call rate below 65% were removed; the same was then done at the sample level. Finally, call rate was recalculated for both analytes and samples and a more stringent 85% threshold was used to filter analytes and samples. After these QC measures,cohorts based on the 7k platform retained aptamer counts of approximately 7,000 for CSF and 6,900 for plasma, whereas cohorts measured on the 5k platform yielded fewer analytes (∼4,700–4,800 unique analytes).

To accommodate variations between the 5k and 7k panels for the CSF data, we restricted analyses to the set of shared analytes in each dataset. For instance, in the internal CSF set, Stanford ADRC (4,735 analytes) and PPMI (4,788 analytes) were combined with cohorts measuring up to 7,075 analytes, yielding an intersection of 3,656 common features (**Supplementary Fig. 2**). We again applied a call-rate threshold of 0.85 to exclude analytes lacking sufficient coverage, ultimately retaining 3,622 CSF analytes for downstream analyses. Meanwhile, the internal plasma dataset uniformly employed SomaScan 7k, beginning with 7,182 analytes; after applying the 85% call-rate filter across cohorts, 6,607 remained for subsequent analysis. A similar approach was applied to merge external cohorts, ensuring that only cross-sectional samples meeting call-rate and QC criteria were included.

For the GNPC dataset, the proteomic data, measured in relative fluorescence units (RFU) using the SomaScan platform, initially included 7,584 analytes corresponding to approximately 7,000 unique proteins. SomaLogic performed initial normalization to remove technical variation across and within plates. Further QC was implemented using an internally developed protocol at both the aptamer and sample levels. A similar procedure to our internal pipeline was applied, including interquartile range (IQR)-based outlier detection and separate call rate thresholds of 65% and 85%. Limit-of-detection, scale factor, and coefficient of variation filtering was not applied because the necessary information was not provided. Additionally, CSF samples measured on the SomaScan 5k and 7k panels were processed separately using the same steps, as the two panels contain different sets of aptamers.

While sparse missing data points can be handled by the classifier, it is important to note that fully missing analytes are not supported. If an analyte that is included in the original dataset is removed from a cohort based on QC, the analyte should nonetheless be kept to ensure a full matrix can be used in the classifier.

After QC and call-rate filtering, we performed a log10 transformation of the remaining aptamer intensities, followed by a z-score normalization procedure in each batch or site (for GNPC). This step aimed to further reduce any batch-specific or technical variability that persisted after initial QC. To confirm that the data were well-harmonized across cohorts, we conducted a principal component analysis (PCA) using participants with definitive disease labels (e.g., AD, PD, FTD, DLB, and CO). The PCA scores (**Supplementary Fig. 1B**) indicated an acceptable inter-cohort alignment, supporting the use of this combined dataset for downstream differential abundance analyses and classification modeling.

### CSF and plasma AD core biomarker measurement and AT classification

We quantified key Alzheimer’s disease (AD) biomarkers, including amyloid-β (Aβ42), phosphorylated tau (pTau)-181, and pTau217 in cerebrospinal fluid (CSF) and plasma samples using multiple immunoassay platforms, including Lumipulse G, Innotest, Roche Elecsys, Alamar NULISAseq, and cobas e 601. Each cohort was evaluated individually, following a revised “AT” classification framework to categorize participants according to amyloid (A) and tau (T) status, as previously described^30,75^.

To derive A and T classifications from quantitative Aβ42 and pTau measurements in CSF, we applied Gaussian mixture models (GMM), which distinguish high versus low levels^61^. Individuals with low CSF Aβ42 and elevated pTau (A+T+) were deemed to have amyloid and tau pathologies, whereas those with high Aβ42 and low pTau (A−T−) were considered to have minimal plaque and tangle burden (i.e., controls). The A+T+ samples that were diagnosed as AD were included in the CSF training/testing, while the A-T-samples diagnosed as CO were also included to ensure biomarker confirmation was present. Intermediate groups (positive for amyloid or ptau, but not both) were used in subsequent analyses. For plasma analyses, because CSF AT status was not available for all samples, we identified biomarker-confirmed controls as those that were CSF A-T-, amyloid PET-, or plasma pTau217^39^. AD cases, on the other hand, were required to be positive for at least one of those biomarkers to be included in the training/testing datasets.

### Differential abundance analysis

Differential abundance analysis was performed using a linear regression model of the formula separately for each fluid:

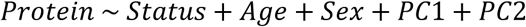

Here, clinical diagnosis (cases vs. controls) was treated as the primary exposure, and age, sex, and the first two principal components (derived via the R package *prcomp*) were included as covariates to account for other biological signals. This model was applied across all disease groups (AD, PD, FTD, DLB). All proteins meeting nominal significance in any of these analyses were combined for subsequent downstream filtering. Analyses were performed by comparing cases to control samples, so the same controls were used for each of the four comparisons. Proteins associated with any disease at a nominal significance threshold were carried forward for subsequent filtering steps.

To eliminate features with minimal variability, we applied a variance threshold of 0.01 using the *VarianceThreshold* function (scikit-learn, Python library), retaining proteins most likely to capture meaningful biological differences.

### Statistical Feature Selection (F-test)

Following the variance threshold step, we employed an F-test to identify the most discriminative features across multiple disease categories. Specifically, we used the *SelectKBest* procedure from scikit-learn with *f_classif* as the scoring function, which calculates the ANOVA F-value for each protein to quantify the ratio of between-group variance to within-group variance. Higher F-values thus indicate stronger discriminatory power for distinguishing among the four neurodegenerative disease groups and healthy controls.

In our experiments, we found that selecting 400 features for CSF data and 700 for plasma data provided optimal performance in terms of macro AUC and accuracy. Adding more features beyond these thresholds did not yield any notable improvement, suggesting that further expansion of the feature space only added noise without enhancing classification results. This F-test-based approach thus helped us maintain a focused set of highly discriminative proteins while ensuring efficient computational performance for subsequent modeling.

### OverSampling Strategy

To address class imbalance—particularly for underrepresented diagnoses such as FTD and DLB, we employed a hybrid oversampling method that combines the Synthetic Minority Over-sampling Technique (SMOTE)^76^ with Tomek link removal. SMOTE artificially synthesizes new minority-class samples by interpolating feature values between existing samples, thereby increasing the representation of minority classes in the training set. Tomek link removal then eliminates ambiguous samples found on the decision boundary between classes, further enhancing class separation and mitigating the risk of introducing noise. This two-stage process aims to improve model performance on rarer disease categories while preserving the overall quality of the training data.

### Multi-Class Classification Model

Based on our pure proteomics data (no covariates incorporated), we implemented a 70% training and 30% testing split using the internal data from the Knight-ADRC, ADNI, PPMI, Barcelona-1, ACE, and Stanford-ADRC, where each disease status was split using the 70-30 ratio to preserve class distributions in both training and testing sets. For model development, we employed a Light Gradient Boosting Machine (LightGBM)^31^, a gradient-boosting framework that constructs an ensemble of weak decision-tree learners. Gradient boosting iteratively improves predictive performance by assigning greater weight to samples that were poorly predicted in earlier iterations, through repeatedly fitting each new tree to the negative gradient (residuals) of the current ensemble and down-weighting well-predicted instances, thereby minimizing a specified loss function. In our multi-class setting, LightGBM produces a five-dimensional probability vector

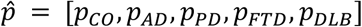

for each sample. Here,

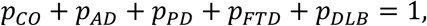

indicating the predicted probability distribution over the five diagnostic categories (healthy controls, AD, PD, FTD, and DLB). These probabilities are obtained using the model’s built-in *predict_proba* method, and the predicted class label is determined as the category with the highest probability.

For model training, we used a “multiclass” objective function combined with the “*multi_logloss*” metric, which measures the divergence between predicted probabilities and ground-truth labels. Specifically, for 𝑁 samples and 𝐾 classes, the multi-class log loss is given by

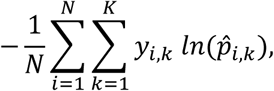

where 𝑦_𝑖,𝑘_ ∈ {0,1} is the true label for class 𝑘 of sample 𝑖, and *p_i,k_* is the predicted probability of class 𝑘.

We determined the number of boosting iterations, 𝑛_𝑒𝑠𝑡𝑖𝑚𝑎𝑡𝑜𝑟𝑠_, empirically to balance model complexity and overfitting. Specifically, we set 𝑛_𝑒𝑠𝑡𝑖𝑚𝑎𝑡𝑜𝑟𝑠_=300 for the CSF dataset and 𝑛_𝑒𝑠𝑡𝑖𝑚𝑎𝑡𝑜𝑟𝑠_=400 for the plasma dataset based on internal validation experiments that determined those values maximized model performance. Additional hyperparameters included a maximum tree depth (*max_depth*=20) to capture complex interactions, a learning rate (*η*=0.05) to moderate incremental updates, and a minimum child-sample threshold (*min_child_samples*=10) to ensure adequate data coverage at each leaf node. The random seed (*random_state*=42) was fixed for reproducibility.

### SHAP for Model Interpretability and Feature-level Attribution

To quantify the contribution of each protein analyte to the LightGBM multi-class classifier, we applied SHapley Additive exPlanations^60^ (SHAP). For a given sample, 𝑥, analyte 𝑖 (out of 𝐹 analytes) and diagnostic class 𝑘 (𝑘 = 1,2, …, 𝐾), the Shapley value is defined:

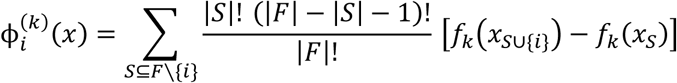

represents the marginal change in the model output 𝑓_𝑘_(𝑥) attributable to adding analyte 𝑖 to every possible combination𝑆 of the remaining analytes. For the held-out test set (𝑁 samples), SHAP returns a three-dimensional tensor,

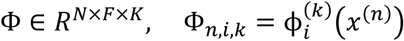

whose *k*-th slice 𝜑_𝑛,𝑖,𝑘_ is visualized as a beeswarm summary plot for class *k*. (**Supplementary Figs. 6 & 7**)

Overall analyte importance was defined as the mean absolute Shapley value across both samples and classes:

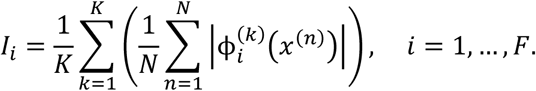

Analytes were ranked by 𝐼_𝑖_ and reported in **Supplementary Tables 7 & 8**.

Beeswarm plots were generated for the top 20 features per class (**Supplementary Figs. 6 & 7**). All analyses were executed under Python 3.11, LightGBM 4.3.0 and SHAP 0.46.0 with a fixed random seed of 42. The complete SHAP tensor and feature ranking tables are available as Supplementary Data files.

### Binary evaluation of a five-class classifier

To enable a direct comparison with binary assays like pTau217 and amyloid-PET, we re-expressed the output of our five-way LightGBM model (classes = AD, CO, DLB, FTD, PD) as a single case-control decision. We converted our multi-disease probabilities to a binary classifier. To do this, we assigned each sample as specific to the class of interest for which it had the highest probability and assigned all other samples with any other class as nonspecific samples. The binary classification was then considered as a one class vs all others calculation. For example, for AD, all samples that had the highest probability across the five classifications for AD were classified as AD, while any sample that was assigned to PD, FTD, DLB, or CO was considered as non-AD. Model performance was then calculated based on a one-vs-all binary classification paradigm using typical metrics (AUC, accuracy, precision, etc.).

### Biomarker Benchmark Logistic Regression Model

To compare our model’s performance to a gold-standard neurodegenerative disease biomarker, we implemented a benchmark logistic regression model to classify AD status using three predictor variables: Age, Sex, and pTau217 & amyloid PET imaging. The dataset was filtered to include only samples with disease status labeled as CO or AD. Additionally, rows with missing values for the predictor variables were removed to ensure complete case analysis.

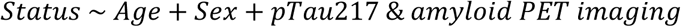

Sex was encoded as a binary variable where Female was mapped to 0 and Male was mapped to 1. The categorical variable T1_pTau217 was transformed into a binary indicator variable with T-coded as 0 and T+ coded as 1.

The dataset was split into training and testing sets with a 70% / 30% ratio using stratified random sampling to preserve the distribution of the CO and AD classes in both subsets. To enable a fair, head-to-head comparison, a fixed random seed was used to control for variability in the data split. Logistic regression was implemented using the *LogisticRegression* function from the *scikit-learn* library, with a maximum iteration threshold of 1000 to ensure model convergence.

Performance metrics, including accuracy, balanced accuracy, macro F1-score, and area under the receiver operating characteristic curve (AUC), were computed for both LightGBM-based and logistic regression-based models. ROC curves were plotted to visualize the classification performance, particularly focusing on the testing set in the 70/30 split model.

### Statistical Analysis

We assessed the concordance between the model’s output probabilities and neuropathological, imaging, clinical, and cognitive assessments of patients who donated samples that were used in this project. For continuous and semi-continuous traits (MMSE, cognitive scores, neuritic plaque burden, neurofibrillary tangle burden, amyloid score, etc.), Pearson correlations between AD & CO probabilities and the phenotype of interest were calculated using the cor.test() function in R. For approximately categorical traits (Braak stage, C score, cognitive diagnosis, PD diagnosis), differences in AD, CO, or PD probability between groups were calculated in two-group comparisons using the Wilcoxon sign-rank test (wilcox.test in R) or across three or more groups using the Kruskal-Wallis test (kruskal.test in R) due to the non-normality of the disease probability distributions. When comparing the distributions of Braak, Global Parkinson’s Score, neurofibrillary tangle burden, and neuritic plaque burden between CO-classified and AD-classified samples, Welch’s t-tests (SciPy, ttest_ind, equal_var = F) were used to compare between the groups because the phenotypes demostrated approximately normal distributions.

For normally distributed variables, including motor-function composites (*motor_gait*), cognitive-domain scores (*cogn_ps*, *cogn_wo*), neuritic-plaque count, neurofibrillary-tangle burden, and Braak stage, two-sided Pearson correlations between every probability and the trait of interest were obtained with **SciPy**’s scipy.stats.pearsonr. Linear trend lines were drawn with seaborn.regplot, omitting scatter points to minimise over-plotting and coloured consistently across all figures (AD = blue, CO = red, FTD = purple, DLB = green, PD = orange). Y-axes were fixed to the theoretical probability range (0–1); x-axis labels reflected the pathology measure analysed. Legends were generated from custom matplotlib.lines.Line2D handles.

Ordinal clinical scales such as *r_pd* (Parkinson’s rating: Not Present → Probable), *dcfdx* (cognitive diagnosis), and other multi-level staging variables were compared across probabilities with non-parametric tests because probability distributions were non-normal. For traits with more than two ordered levels, an omnibus Kruskal–Wallis H-test (scipy.stats.kruskal) was applied; significant findings triggered pairwise Wilcoxon rank-sum tests (scipy.stats.ranksums). For binary traits with independent groups (e.g., CDR=0 vs. CDR≥1), Wilcoxon rank-sum tests were used directly. Group means ± s.e.m. were depicted using **seaborn** barplots (errorbar =’se’, estimator =’mean’), colour-coded consistently for the five predicted classes. Progression from CO to AD was compared between individuals assigned high probability of AD (AD probability ≥ 80%) and individuals assigned high probability of CO (CO probability ≥ 80%) using a Cox Regression model, adjusted for age at sample draw. Because exact diagnosis date was not available, the time span between sample draw and diagnosis was set as the time between sample draw and final diagnosis for each sample. Models were calculated using the coxph command in the survival R package (v3.7-0). To make the Kaplan-Meier plots, data from each fluid was first fit to a survival model using the survfit command in the survival R package. Plots were then generated using the ggsurvplot command in the survminer R package (v0.5.0).

All analyses were executed in Python 3.10.13 with pandas 2.2.2, SciPy 1.11.4, Matplotlib 3.8.4 and Seaborn 0.13.2 or R v4.4.0.

## Acknowledgements

We extend our gratitude to all the participants, their families, as well as the cohorts, institutions, and their dedicated staff.

Funding: This work was supported by grants from the National Institutes of Health (R01AG044546 (CC), P30AG066444 (DMH), RF1AG053303 (CC), RF1AG058501 (CC), U01AG058922 (CC), the Chan Zuckerberg Initiative (CZI), the Michael J. Fox Foundation (CC), the Alzheimer’s Association Zenith Fellows Award (ZEN-22-848604, awarded to CC), and an Anonymous foundation. A.S. receives support from multiple NIH grants (P30 AG010133, P30 AG072976, R01 AG075959, R01 AG082348, R01 AG081951, R01 AG057739, R01 AG070883, U01 AG024904, R01 LM013463, T32 AG071444, U24 AG074855, U01 AG068057, U01 AG072177, U01 AG24904, U19 AG074879, U19 AG024904, R01 LM013463, and R01 AG068193). K.N. receives support from NIH grants (R01 AG081951, R01 LM012535, U01 AG072177, and U19 AG074879).

This work was supported by access to equipment made possible by the Hope Center for Neurological Disorders, the Neurogenomics and Informatics Center (NGI: https://neurogenomics.wustl.edu/) and the Departments of Neurology and Psychiatry at Washington University School of Medicine.

DIAN resources: Data collection and sharing for this project was supported by The Dominantly Inherited Alzheimer Network (DIAN, U19AG032438) funded by the National Institute on Aging (NIA), the Alzheimer’s Association (SG-20-690363-DIAN).

ACE Alzheimer Center: Authors acknowledge the support of the Agency for Innovation and Entrepreneurship (VLAIO) grant No PR067/21 for the HARPONE project. Also, the Spanish Ministry of Science and Innovation, Proyectos de Generación de Conocimiento grant PID2021-122473OA-I00, Instituto de Salud Carlos III (ISCIII), Acción Estratégica en Salud integrated in the Spanish National R+D+I Plan and financed by ISCIII Subdirección General de Evaluación and the Fondo Europeo de Desarrollo Regional (FEDER “Una manera de hacer Europa”) grants PI19/00335 and PI22/01403. The support of CIBERNED (ISCIII) under the grant CB18/05/00010. The support from PREADAPT project, Joint Program for Neurodegenerative Diseases (JPND) grant No AC19/00097. The support of Fundación bancaria “La Caixa”, Fundación ADEY, Fundación Echevarne and Grífols SA (GR@ACE project). A.C. received support from the ISCIII under the grant Sara Borrell (CD22/00125). P.G.G. is supported by CIBERNED employment plan (CNV-304-PRF-866). AR is also supported by STAR Award. University of Texas System. Tx, United States, The South Texas ADRC. National Institute of Aging. National Institutes of Heath. USA. (P30AG066546), the Keith M. Orme and Pat Vigeon Orme Endowed Chair in Alzheimer’s and Neurodegenerative Diseases and Patricia Ruth Frederick Distinguished Chair for Precision Therapeutics in Alzheimer’s and Neurodegenerative Diseases.

## Data availability

Proteomic data can be requested for the following cohorts: ADNI (https://adni.loni.usc.edu/), Knight-ADRC (https://knightadrc.wustl.edu/data-request-form/), Stanford-ADRC (https://med.stanford.edu/adrc/researcher-resources.html), DIAN (https://dian.wustl.edu/our-research/for-investigators/diantu-investigator-resources/dian-tu-biospecimen-request-form/), PPMI (https://www.ppmi-info.org/access-data-specimens/download-data), ROSMAP (https://www.radc.rush.edu/), and the I-ADRC (https://medicine.iu.edu/research-centers/alzheimers/for-researchers/resource-request). Proteomic data from the GNPC used in the preparation of this manuscript will be shared through the GNPC website (ttps://www.neuroproteome.org/) in July 2025.

Due to GDPR restrictions, data from the Barcelona-1 and ACE cohorts are not publicly available. Participants from the WashU Movement Disorder Clinic have not given consent for individual-level data sharing.

## Code availability

Code for our classifier development, evaluation, & neuropathological trail and cognitive tests statistical correlation all can be found at https://github.com/NeuroGenomicsAndInformatics/2025_Xu_CSF_Plasma_AI_Classifier

