## Supplementary Figure for "Protein-based Diagnosis and Analysis of Co-pathologies Across Neurodegenerative Diseases: Large-Scale AI-Boosted CSF and Plasma Classification"

**CSF and Plasma Classifier Supplementary Figures**

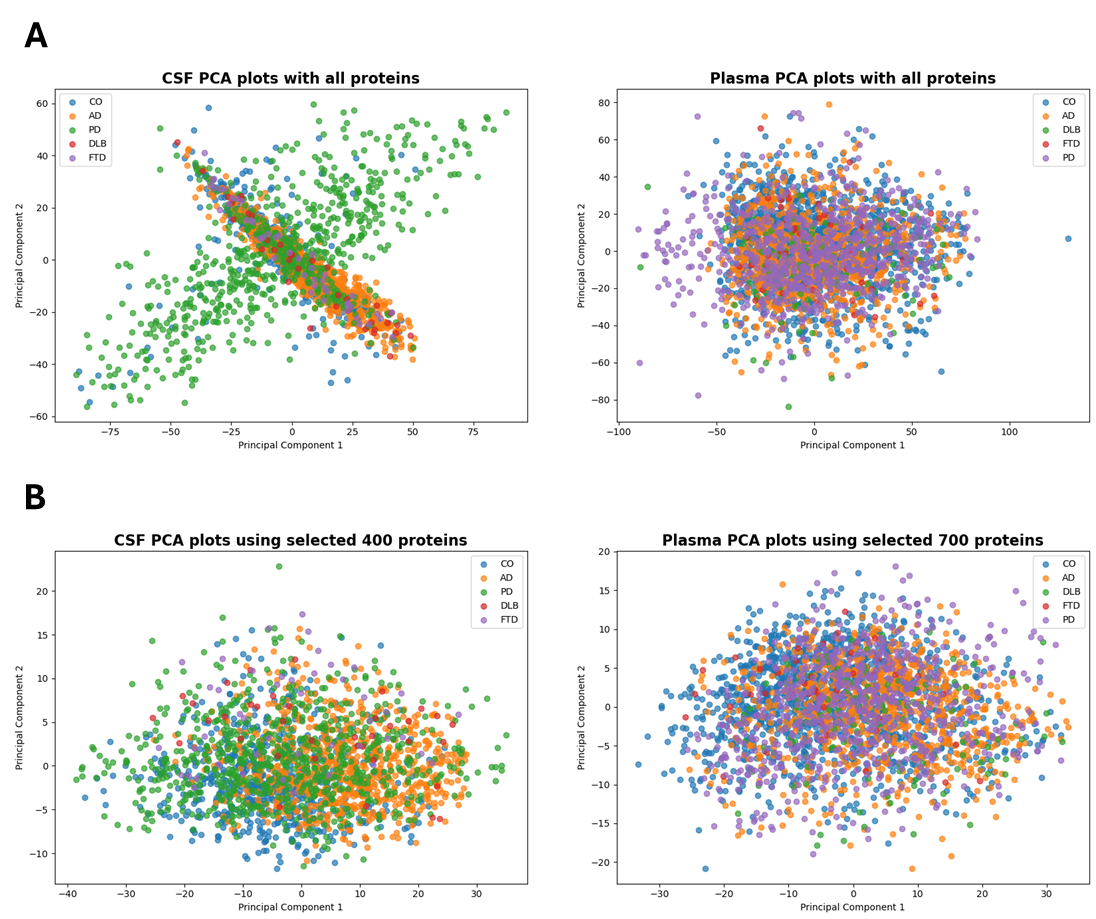
**Supplementary Figure 1: Principal Component Analysis (PCA) of CSF and Plasma proteomic profiles. (A) PCA plots using all detected proteins in cerebrospinal fluid (CSF; left) and plasma (right).** Samples are color-coded by clinical diagnosis: CO, AD, PD, FTD, DLB. **(B) PCA plots using selected proteins (n=400 for CSF, n=700 for plasma) identified through feature selection.** The reduced feature sets improve group separation in CSF (left) while showing limited separation in plasma (right). Colors correspond to diagnostic categories.

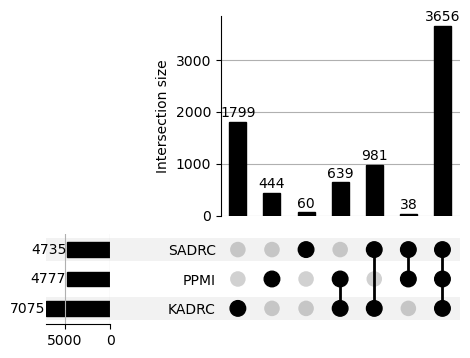

**Supplementary Figure 2: CSF overlapping analyte selection.** UpSet plot showing the intersection of analytes detected in cerebrospinal fluid (CSF) across three cohorts: SADRC, PPMI, and KADRC. Each horizontal bar on the left indicates the total number of analytes measured in each cohort (4735 for SADRC (SomaScan 5k), 4777 for PPMI (custom SomaScan 5k), and 7075 for KADRC(SomaScan 7k)). The vertical bars represent the size of the intersection across different cohort combinations, with black dots and connecting lines below indicating the cohort(s) included in each intersection set. The largest shared subset (n = 3656) was common to all three cohorts. These analytes were carried forward for further filtering and analysis in CSF.

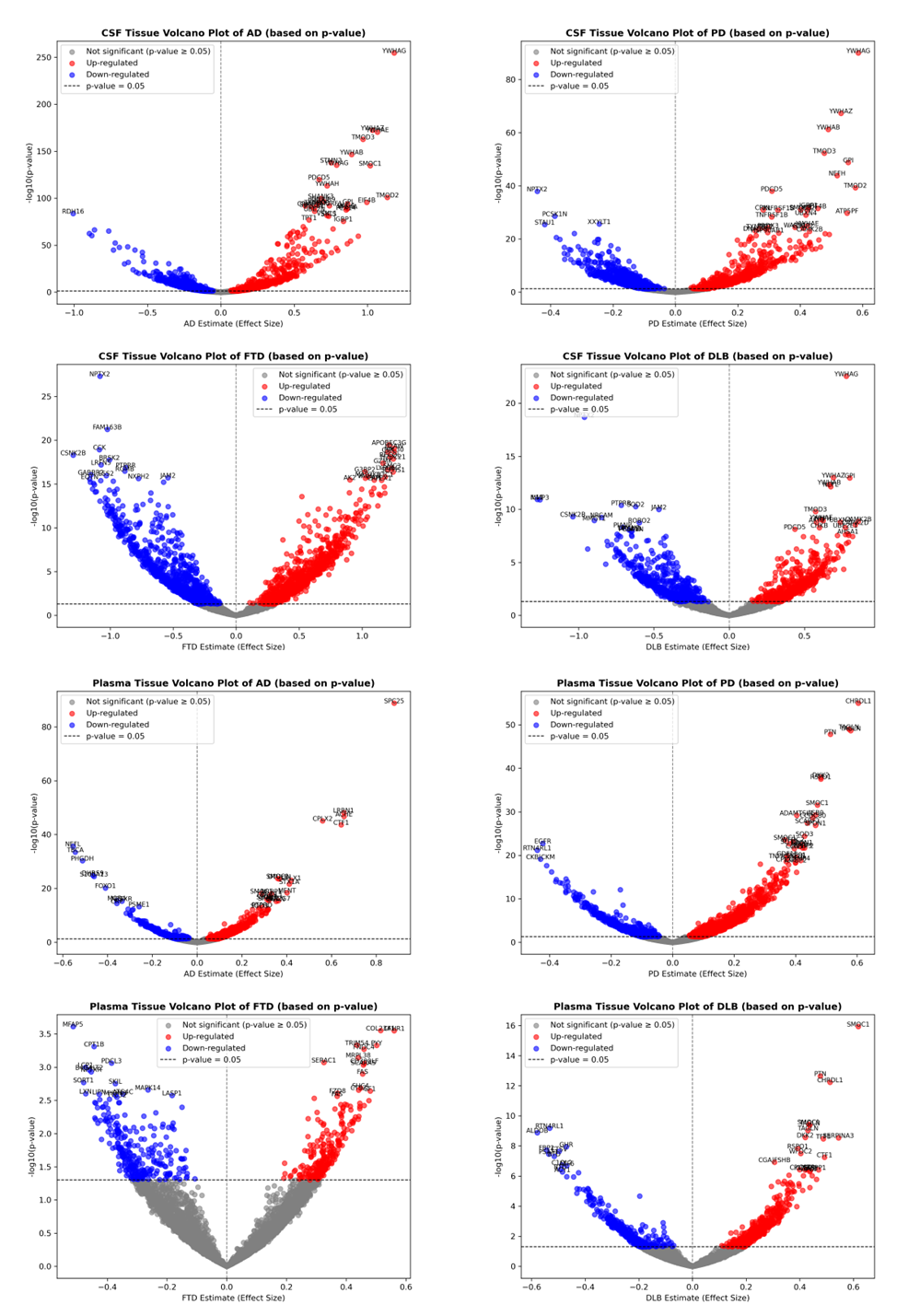
**Supplementary Figure 3: Volcano plots of differentially expressed proteins (DEPs) in CSF and plasma across neurodegenerative diseases (p-value based).** Each panel displays the distribution of DEPs in either cerebrospinal fluid (CSF; top two rows) or plasma (bottom two rows), comparing disease versus control samples. The x-axis represents the estimated effect size (log fold-change), and the y-axis shows the −log10(p-value). Proteins with p-values < 0.05 are color-coded as up-regulated (red) or down-regulated (blue), while non-significant proteins are shown in grey. Top row: CSF volcano plots for Alzheimer’s disease (AD), Parkinson’s disease (PD), frontotemporal dementia (FTD), and dementia with Lewy bodies (DLB). Bottom row: Plasma volcano plots for the same four diseases. Notable up- and down-regulated proteins are annotated with gene names.

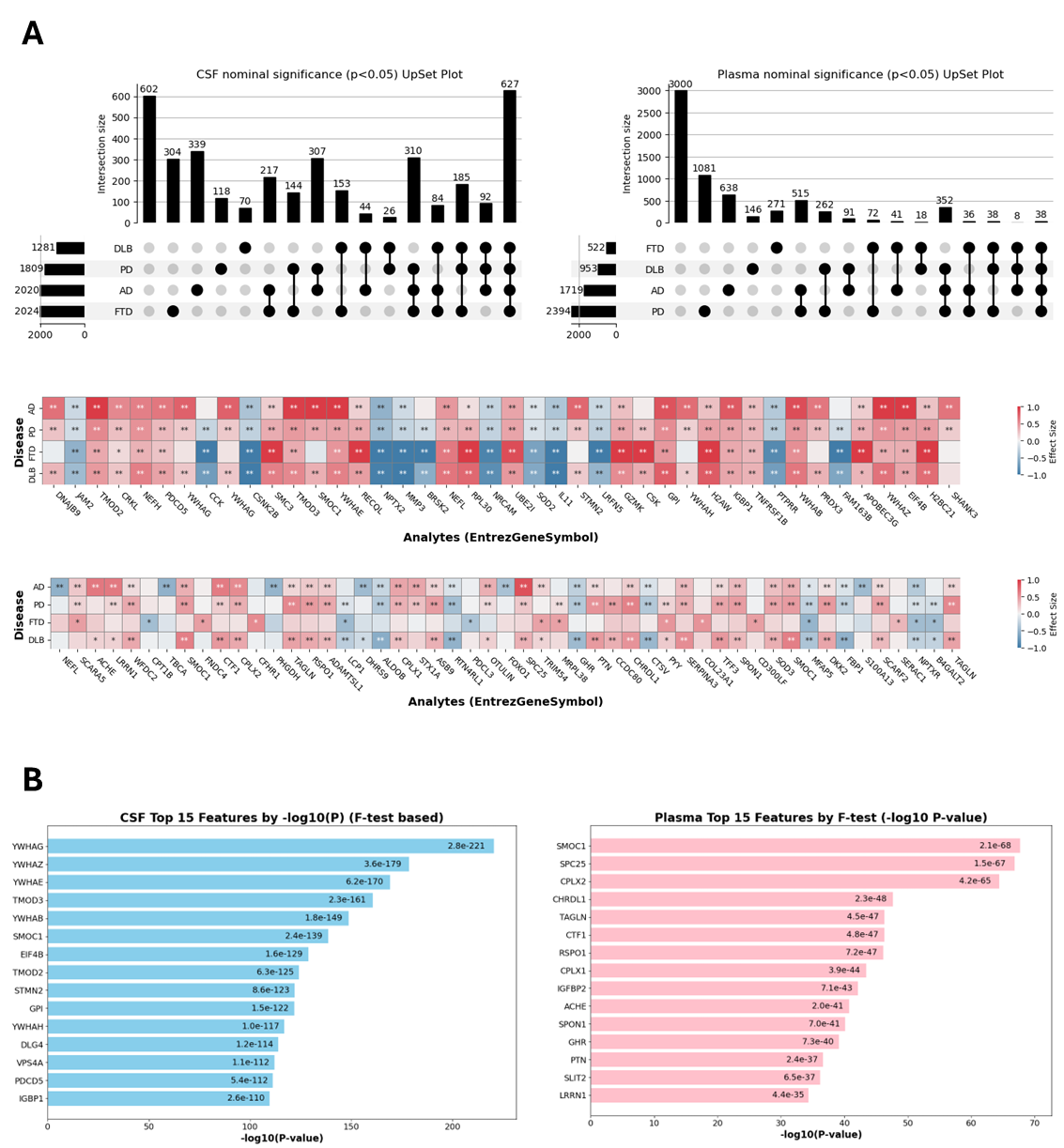
**Supplementary Figure 4. Cross-disease overview of CSF and plasma proteomic signals. (A) Co-expressed proteins.** Top: CSF & Plasma UpSet plot of nominally significant analytes (p < 0.05). Horizontal bars show the total number of significant analytes detected for each disease (AD, PD, DLB, FTD). Vertical bars give the size of every intersection set, with filled circles beneath indicating the disease(s) represented in that intersection. Middle: Heat-maps of the top 15 CSF & plasma analytes ranked by p-value per disease. Rows correspond to the four diseases, and columns to analytes (labeled with EntrezGeneSymbols). Cell color denotes standardized effect size (red = up-regulation, blue = down-regulation; scale at right). Some proteins were in the top 15 for multiple diseases and are therefore listed once, so fewer than 60 total proteins are listed. **(B) F-test based single-analyte p-values.** Bar chart of the 15 most informative CSF (left) & plasma (right) features selected by F-test. Bars are sorted by −log10(p-value) on a log scale; labels show the exact p-value.

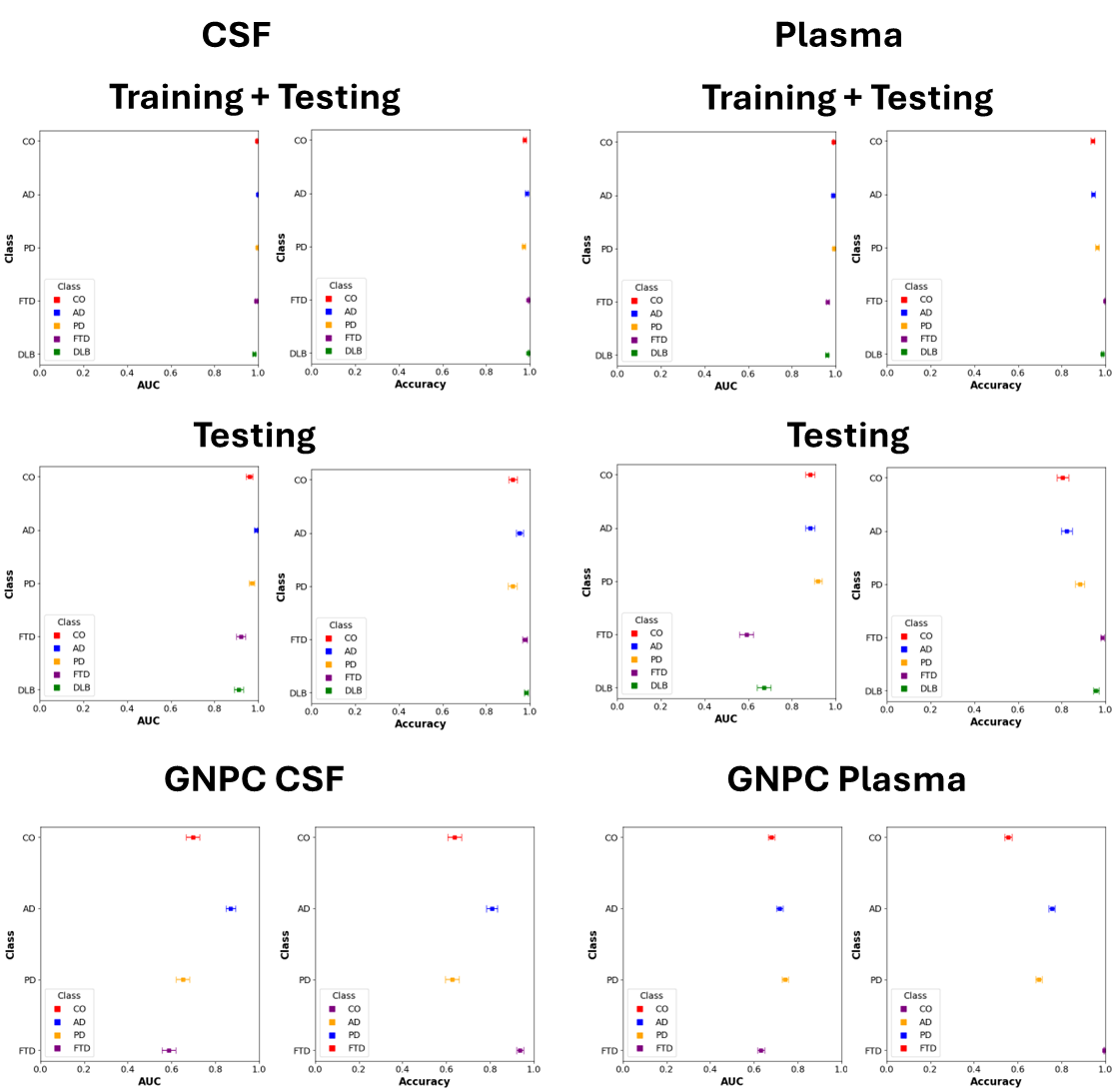
**Figure 5. Diagnostic performance of the five-class proteomic classifier across fluids and evaluation cohorts. C**lass-specific performance of models trained on cerebrospinal fluid (CSF, left panels) and plasma (right panels). Top panels: Whisker plots for Area under the Receiver-Operator Curve (AUC) and accuracy for combined training & testing datasets. Middle panels: Whisker plots for AUC and accuracy for testing dataset only. Bottom panels: Whisker plots for AUC and accuracy for the GNPC dataset. Dot color denotes sample status (CO, red; AD, blue; PD, yellow; FTD, purple; DLB, green). Horizontal whiskers represent 95 % confidence intervals.

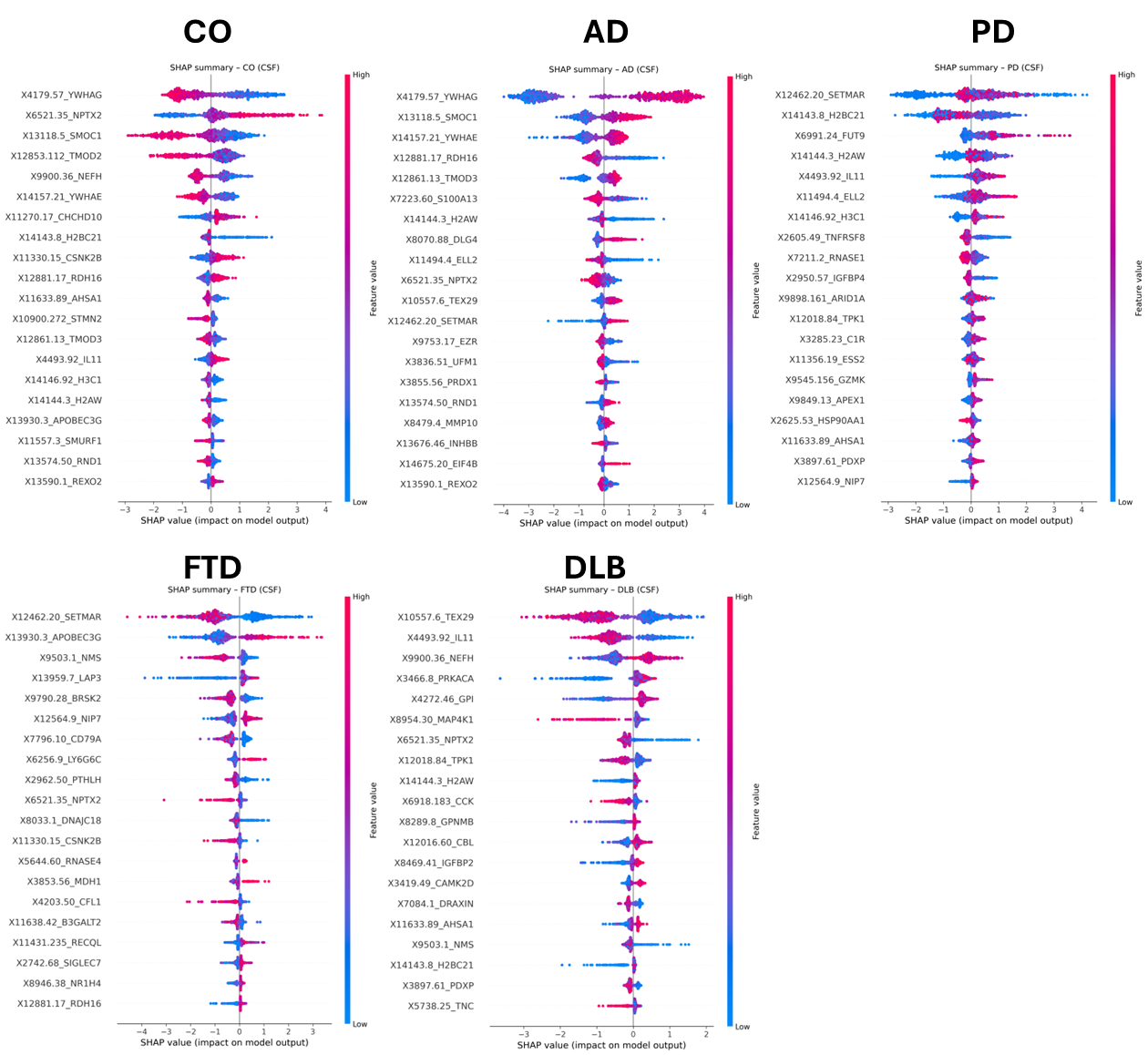
**Supplementary Figure 6. SHAP summary plots for the CSF‐based classifier.** SHAP (SHapley Additive exPlanations) bee-swarm plots display the 20 most influential CSF proteins for each diagnostic class: CO, AD, PD, FTD, and DLB (left-to-right, top row; left-to-right, bottom row). Each dot represents a single participant and is positioned on the *x*-axis by the magnitude and direction of that protein’s contribution to the LightGBM model’s log-odds for the indicated class (positive values push the prediction toward the class, negative values away). Dots are color-coded by *z*-scored protein abundance (blue = low, pink = high), illustrating whether increased or decreased levels drive the class-specific prediction. Protein identifiers are reported with EntrezGeneSymbols.

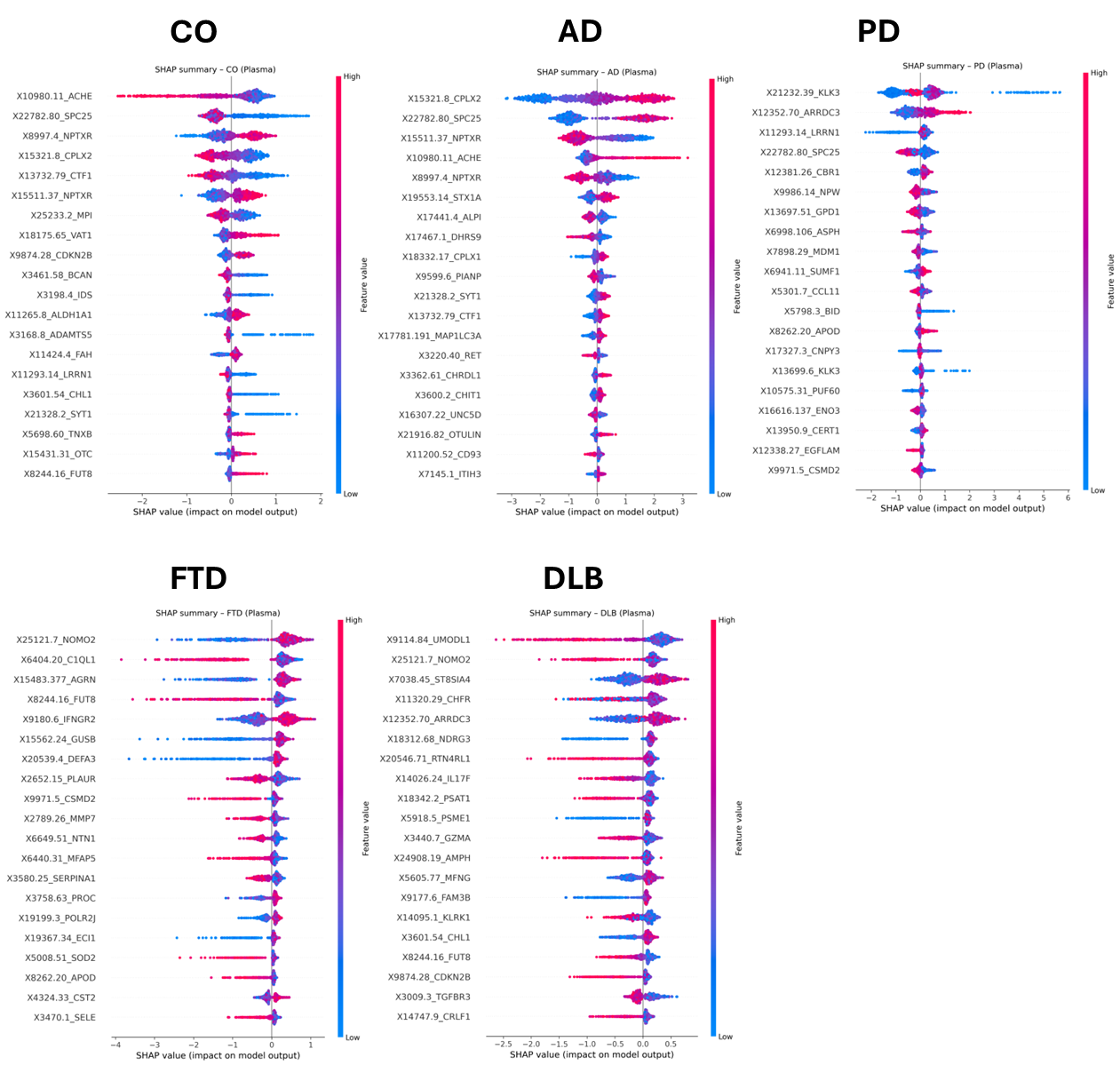
**Supplementary Figure 7. SHAP summary plots for the plasma-based classifier.** Analogous SHAP bee-swarm plots for the plasma model that include the top 20 plasma proteins most influential for distinguishing CO, AD, PD, FTD, and DLB. Interpretation is identical to the CSF plots: dot position reflects contribution magnitude and sign, while color denotes standardized protein abundance.

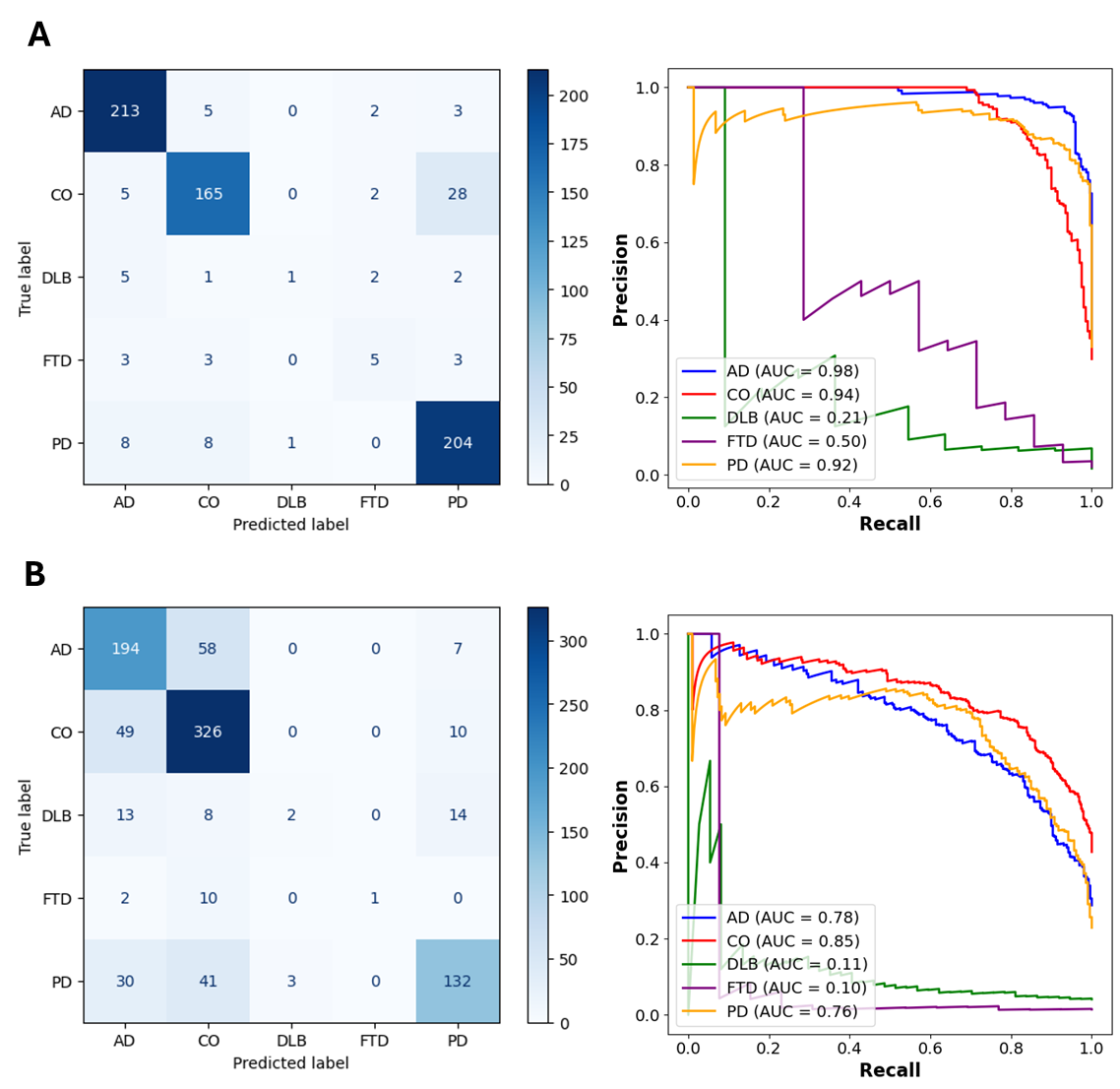
**Supplementary Figure 8. Multi-class disease classification performance using CSF and plasma proteomic profiles. (A) CSF-based model performance in the testing dataset.** Left: Confusion matrix of the CSF-based model evaluated on the testing set, showing true versus predicted diagnostic labels across five classes: Alzheimer’s disease (AD), cognitively normal, biomarker-confirmed controls (CO), dementia with Lewy bodies (DLB), frontotemporal dementia (FTD), and Parkinson’s disease (PD). Precision-recall (PR) curves for each diagnostic class, with AUC values indicated in the legend (macro AUC = 0.71). The macro-averaged AUC across all classes is 0.70. **(B) Plasma-based model performance.** Left: Confusion matrix of the plasma-based model evaluated on the testing set, using the same five-class classification scheme. Right: PR curves for each class (macro AUC = 0.52).

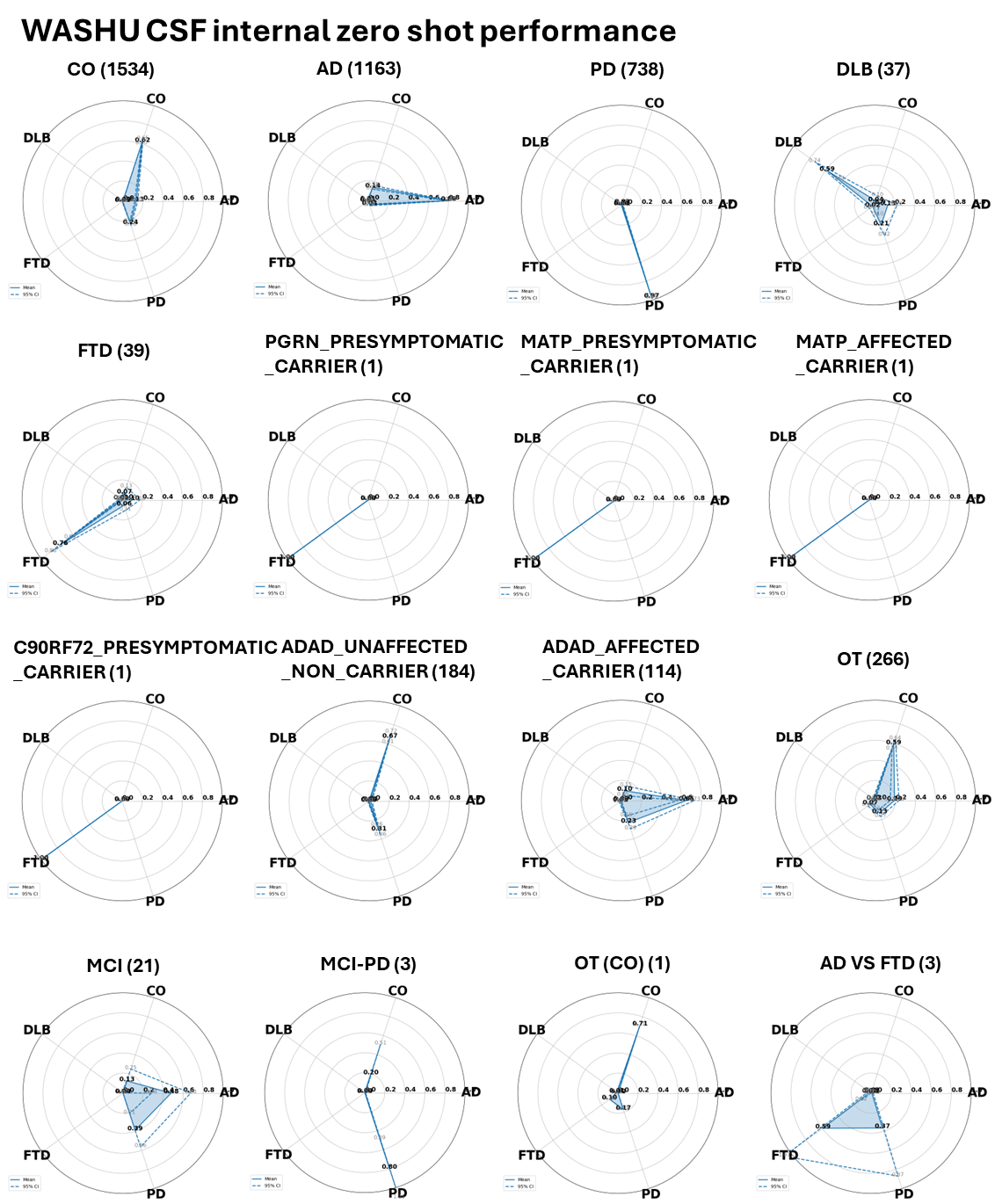

**Supplementary Figure 9. Zero-shot inference by the CSF classifier in the internal cohorts.** Each radar chart summarizes the classifier’s output for a distinct clinical or genetic subgroup that was not used for model training (group name followed by sample size in parentheses). Axes correspond to the five diagnostic logits returned by the model—Alzheimer’s disease (AD), Parkinson’s disease (PD), frontotemporal dementia (FTD), dementia with Lewy bodies (DLB) and cognitively normal controls (CO). Solid blue polygon, mean predicted probability vector across all samples in the subgroup; Dashed line, 95% confidence envelope obtained by bootstrapping individuals within the subgroup. PGRN Presymptomatic Mutation Carrier, carrier of a FTD-causing mutation in PGRN that has not yet developed symptoms; MAPT Presymptomatic Mutation Carrier, carrier of an FTD-causing mutation in MAPT that has not yet developed symptoms; MAPT Affected Carrier, carrier of an FTD-causing mutation in MAPT that has been clinically diagnosed with FTD; C9orf72 Presymptomatic Carrier: carrier of an FTD-causing mutation in C9orf72 that has not yet developed symptoms; DIAN Noncarrier controls: individuals part of the DIAN study of autosomal-dominant AD (ADAD) that do not carry ADAD-causal mutations and have not developed other dementias; ADAD Affected Mutation Carriers, carriers of an ADAD-causing mutation that have been clinically diagnosed with AD; OT: other unspecified form of dementia; MCI, mild cognitive impairment; MCI-PD, MCI & PD; OT (CO), cognitively healthy but too young to be considered a control; AD VS FTD, evidence of both AD & FTD.

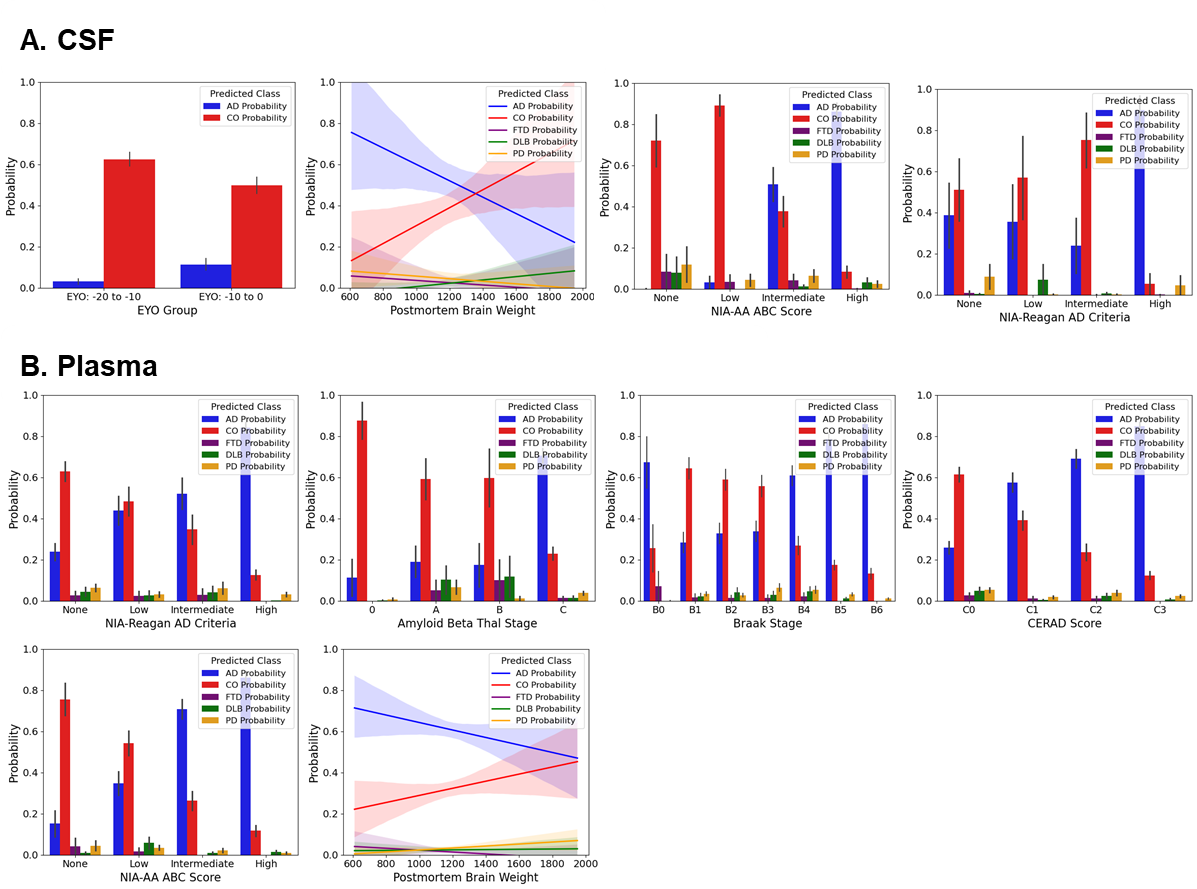
**Supplementary Figure 10. Predicted disease probabilities track clinical and neuropathological severity.** **A, Concordance between CSF-based model probabilities and neuropathological findings or genetic evidence.** Plot 1, barplot of mean AD & CO probabilities within presymptomatic AD mutation carriers from DIAN, separated by the expected years until symptom onset (EYO, ± SE). Plot 2, plot of linear regression of mean probabilities against postmortem brain weight (LOESS curve ± 95% CI). Plot 3, barplot of mean probabilities within different NIA-AA-defined ABC score categories (summary of amyloid, tau, and neuritic plaque presence in the brain, ±SE). Plot 4, barplot of mean probabilities within different NIA-Reagan AD diagnostic criteria groups (±SE). **B, Concordance between plasma-based model probabilities and neuropathological findings.** From top left to bottom right, Plot 1, bar plot of mean probabilities within different groups based on the NIA-Reagan AD criteria (±SE). Plot 2, bar plot of mean probabilities within different groups based on Thal stage (±SE). Plot 3, bar plot of mean probabilities within different groups based on Braak stage (±SE). Plot 4, bar plot of mean probabilities within different groups based on CERAD score (±SE). Plot 5, bar plot of mean probabilities within different groups based on the NIA-AA ABC composite AD score (±SE). Plot 6, plot of linear regression of mean probabilities against postmortem brain weight (LOESS curve ± 95% CI).

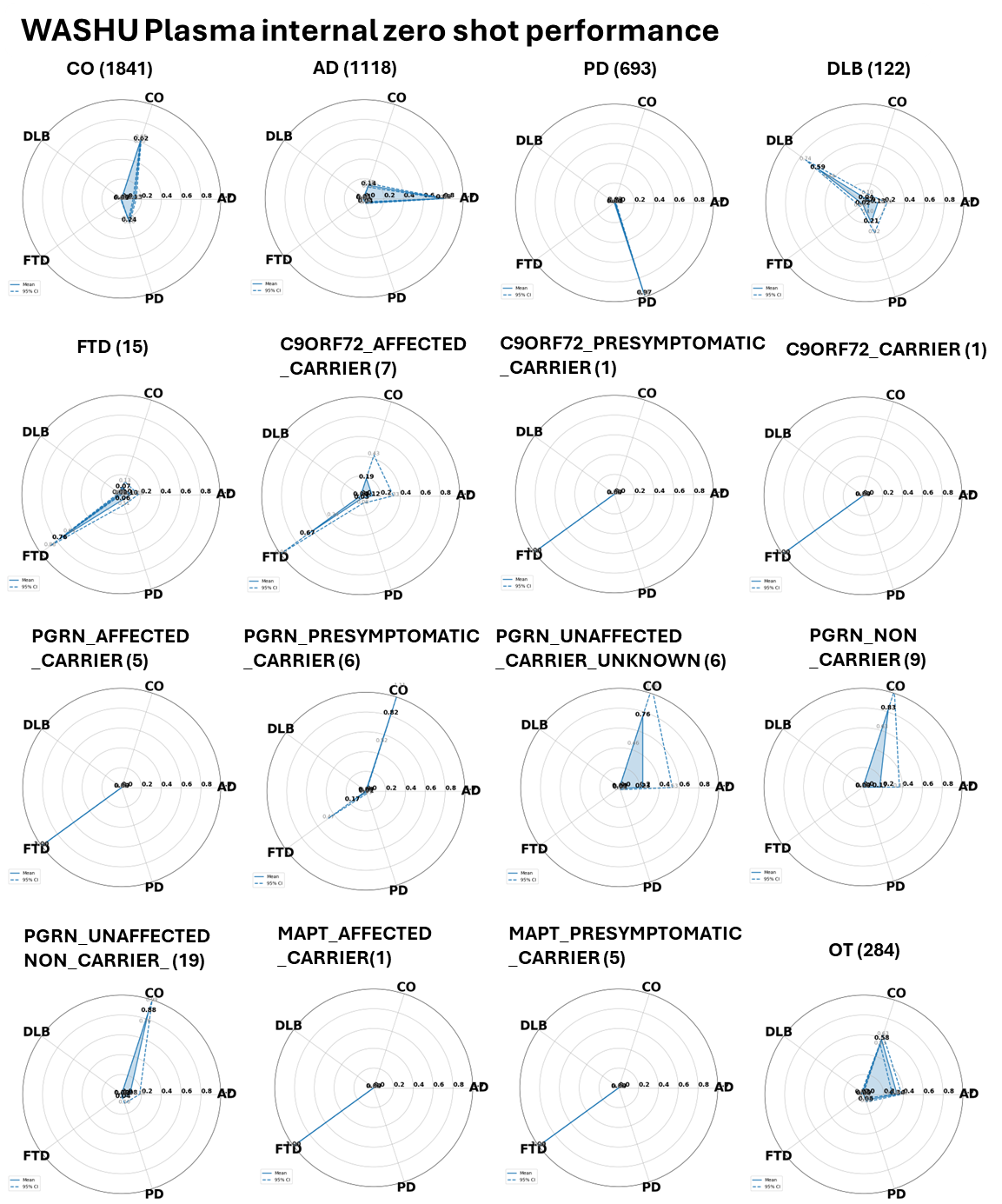

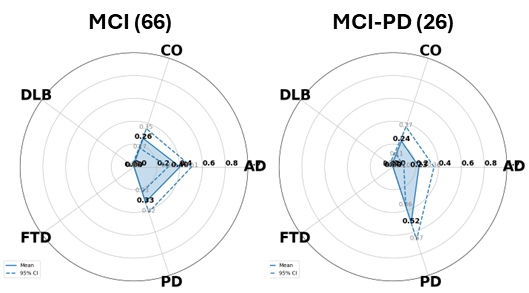

**Supplementary Figure 11. Zero-shot inference by the plasma classifier in the internal cohorts.** Each radar chart summarizes the classifier’s output for a distinct clinical or genetic subgroup that was not used for model training (group name followed by sample size in parentheses). Axes correspond to the five diagnostic logits returned by the model—Alzheimer’s disease (AD), Parkinson’s disease (PD), frontotemporal dementia (FTD), dementia with Lewy bodies (DLB) and cognitively normal controls (CO). Solid blue polygon, mean predicted probability vector across all samples in the subgroup; Dashed line, 95% CI obtained by bootstrapping individuals within the subgroup. C9orf72 Affected Carrier, carrier of an FTD-causing mutation in C9orf72 that is clinically diagnosed with FTD; C9orf72 Presymptomatic Carrier, carrier of an FTD-causing mutation in C9orf72 that has not yet developed FTD symptoms; C9orf72 Carrier: carrier of an FTD-causing mutation in C9orf72 with unknown clinical status; PGRN Affected Carrier, carrier of an FTD-causing mutation in C9orf72 that is clinically diagnosed with FTD; PGRN Presymptomatic Carrier, carrier of a FTD-causing mutation in PGRN that has not yet developed symptoms; PGRN Unaffected Carrier Unknown: individual without symptoms at plasma draw who does not have clear PGRN mutation status; PGRN Noncarrier, individual without an FTD-causing PGRN mutation without known clinical status; PGRN Unaffected Noncarrier, Individual who is known to not carry a PGRN mutation who does not have clinical dementia; MAPT Affected Carrier, carrier of an FTD-causing mutation in MAPT that has been clinically diagnosed with FTD; MAPT Presymptomatic Mutation Carrier, carrier of an FTD-causing mutation in MAPT that has not yet developed symptoms; OT: other unspecified form of dementia; MCI, mild cognitive impairment; MCI-PD, mild cognitive impairment and PD.

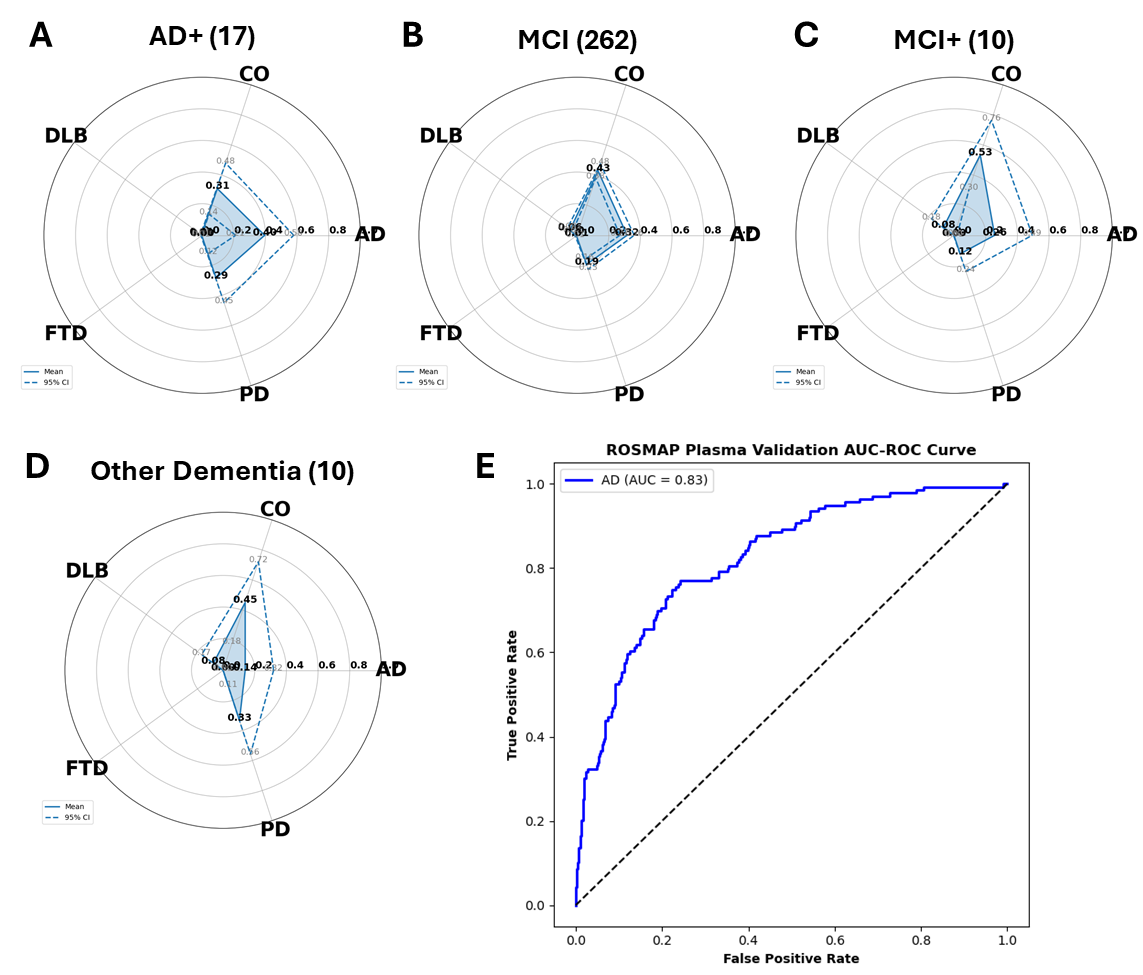
**Supplementary Figure 12. External validation of the plasma-based classifier in ROSMAP.** **(A–D) Group-level probability fingerprints.** Radar plots depict the mean predicted probability (solid line) for each of the five output classes (AD, PD, FTD, DLB, CO) with the 95 % confidence interval. A, AD plus another condition contributing to cognitive impairment); B, Mild Cognitive Impairment; C, Mild cognitive impairment and another condition contributing to cognitive impairment; D, Primary cause of dementia is something other than AD. **(E) Receiver-operating–characteristic analysis.** One-vs-rest ROC curves assess binary discrimination between autopsy-confirmed AD+ cases (blue) or CO cases (orange) and all remaining ROSMAP participants. Areas under the curve are AUC = 0.80 for AD+ and AUC = 0.79 for CO; the dashed diagonal indicates chance level.

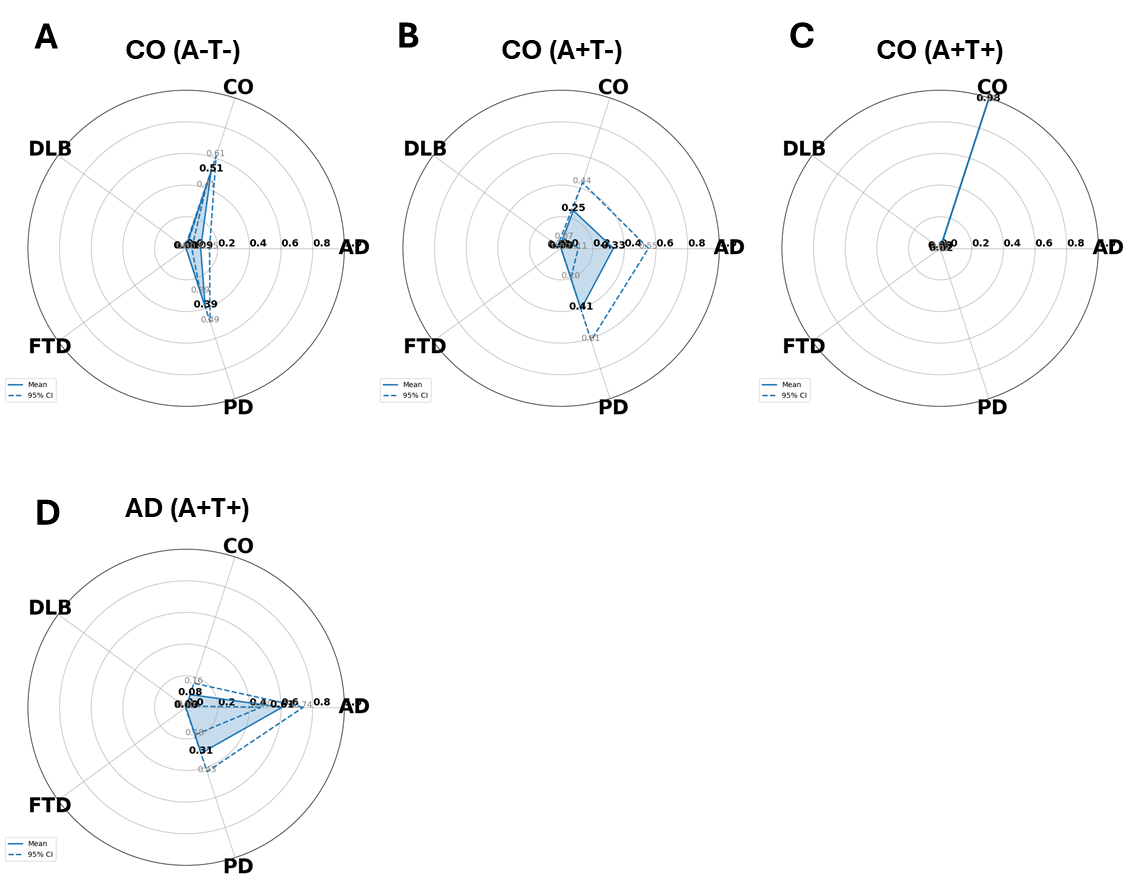
**Supplementary Figure 13. External validation of the plasma-based classifier in the Indiana ADRC.** **(A–D) Group-level probability fingerprints.** **A**, plot for CSF biomarker-confirmed (amyloid and tau negative) cognitively intact controls (A-T-). **B**, plot for cognitively intact controls that are positive for CSF amyloid but negative for CSF tau levels (A+T-). **C,** plot for cognitively intact individuals that are positive for both CSF amyloid and tau (A+T+). **D,** plot for clinically-diagnosed AD individuals who are positive for both CSF amyloid and tau (A+T+). Plots depict the mean predicted probability (solid line) for each of the five output classes, AD, PD, FTD, DLB, and CO, together with the 95% confidence interval.

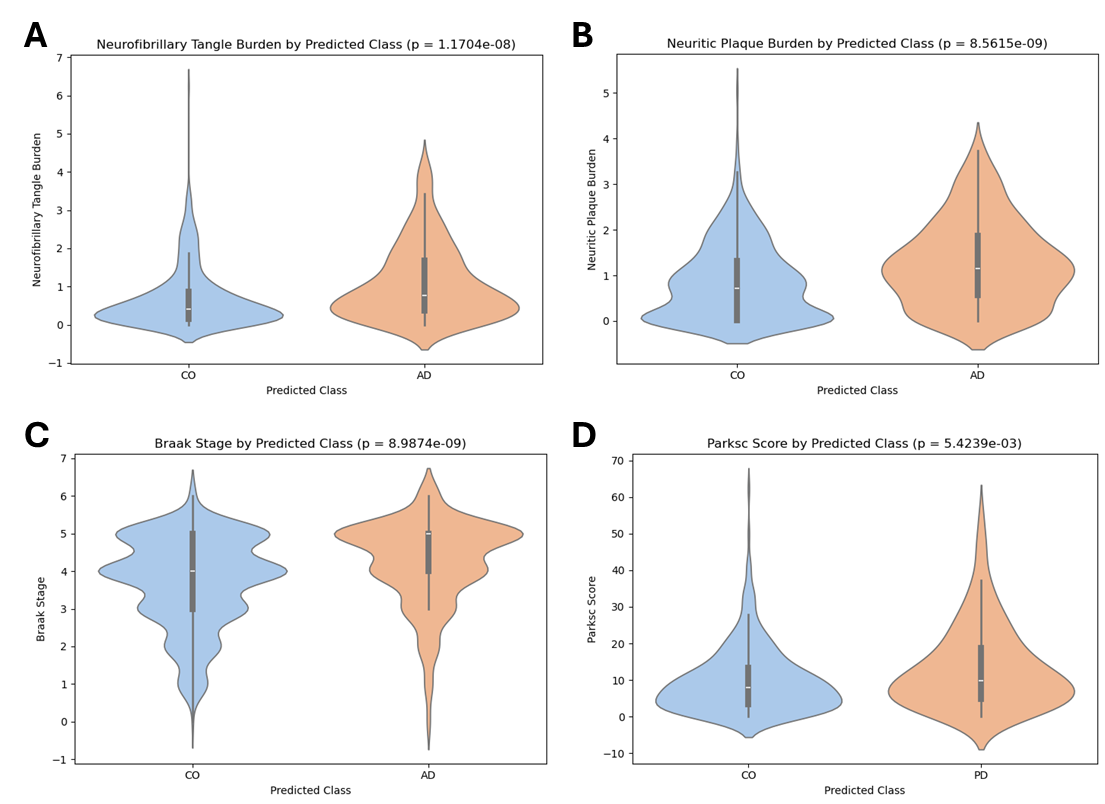
**Supplementary Figure 14. Disease-classified samples from ROSMAP have higher disease burden than CO-classified samples. (A) Neurofibrillary tangle burden.** Predicted AD cases (n=167, orange) display a higher tangle burden than CO (*n* = 507, blue; *P* = 1.17 × 10⁻⁸). **(B) Neuritic plaque burden.** Predicted AD cases have a greater amyloid plaque burden than CO (*P* = 8.56 × 10⁻⁹). **(C) Braak stage.** Individuals predicted as AD show markedly higher Braak stage than those predicted as CO (*P* = 8.99 × 10⁻⁹). **(D) Global Parkinsonian score (parksc).** Samples predicted as Parkinson’s disease (PD, *n* = 167) exhibit significantly greater motor impairment than CO (*n* = 507; *P* = 5.42 × 10⁻³). White tick represents median value; gray bars represent interquartile range. P-values are from two-sided Welch’s *t*-tests and are displayed in panel titles.

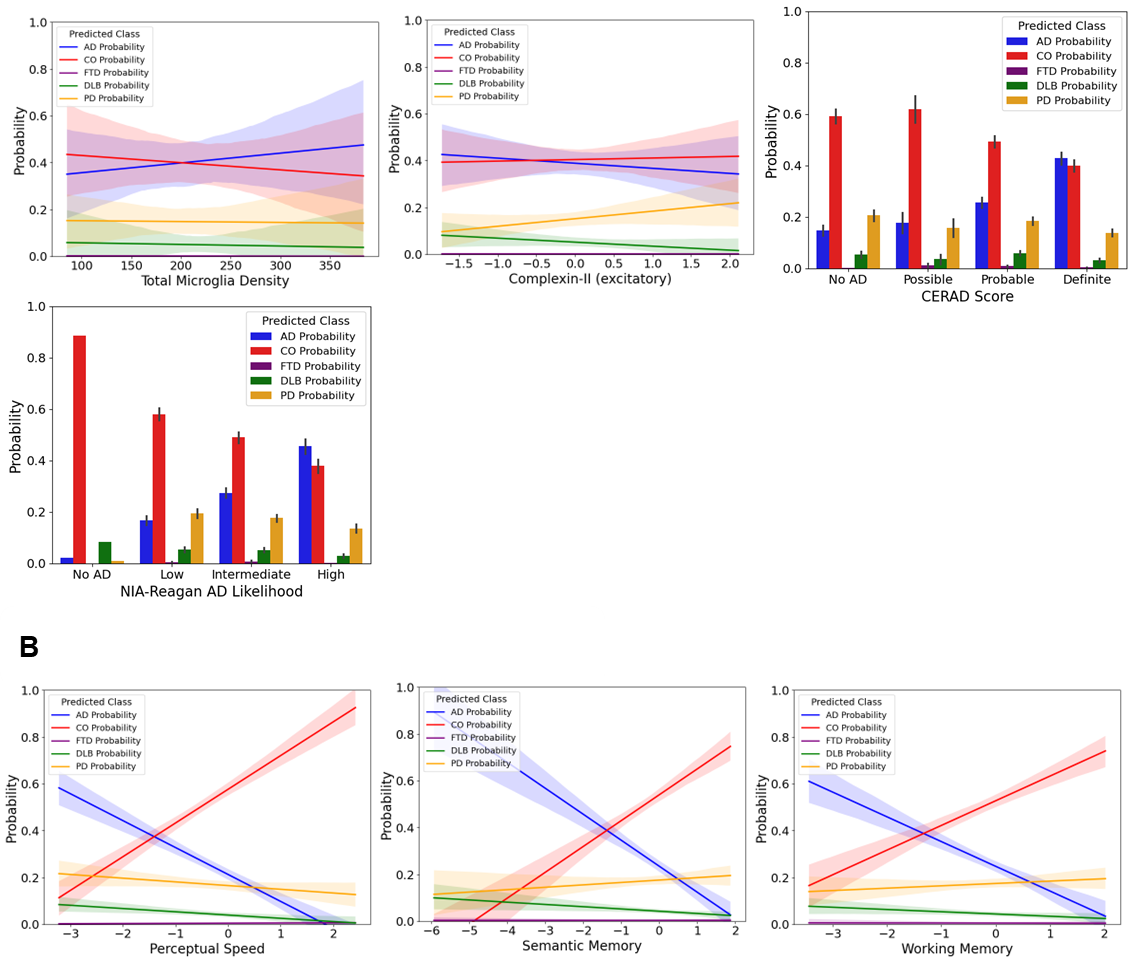
**Supplementary Figure 15. Plasma-derived diagnostic probabilities from the ROSMAP cohort mirror neuropathological burden and cognitive performance. (A) Neuropathological traits.** Plot 1, linear regression of model-defined probabilities for each class against total microglia density in the ventral medial caudate of the brain. Plot 2, linear regression of model probabilities against z-score-normalized mean Complexin-II protein levels across multiple brain regions. Plot 3, bar plot of mean probabilities (±SE) within different groups based on CERAD score, ranging from no AD evidence to definite AD. Plot 4, bar plot of mean probabilities (±SE) within different groups based on NIA-Reagan classification, ranging from no AD evidence to high likelihood of AD. modelled with simple linear regression (solid lines) and 95 % confidence bands (shaded). **(B) Cognitive tests.** Plot 1, linear regression of model-defined probabilities against normalized scores for perceptual speed, a measure of cognitive function. Plot 2, linear regression of model-defined probabilities against normalized scores for semantic memory, a measure of cognitive function related to memory of everyday objects. Plot 3, linear regression of model-defined probabilities against normalized scores for working memory, a measure of cognitive function related to decision making and task completion. Lines represent regression values ± 95% CI. Alzheimer’s disease (AD, blue); cognitively unimpaired controls (CO, red); Parkinson’s disease (PD, orange); dementia with Lewy bodies (DLB, green); frontotemporal dementia (FTD, purple).

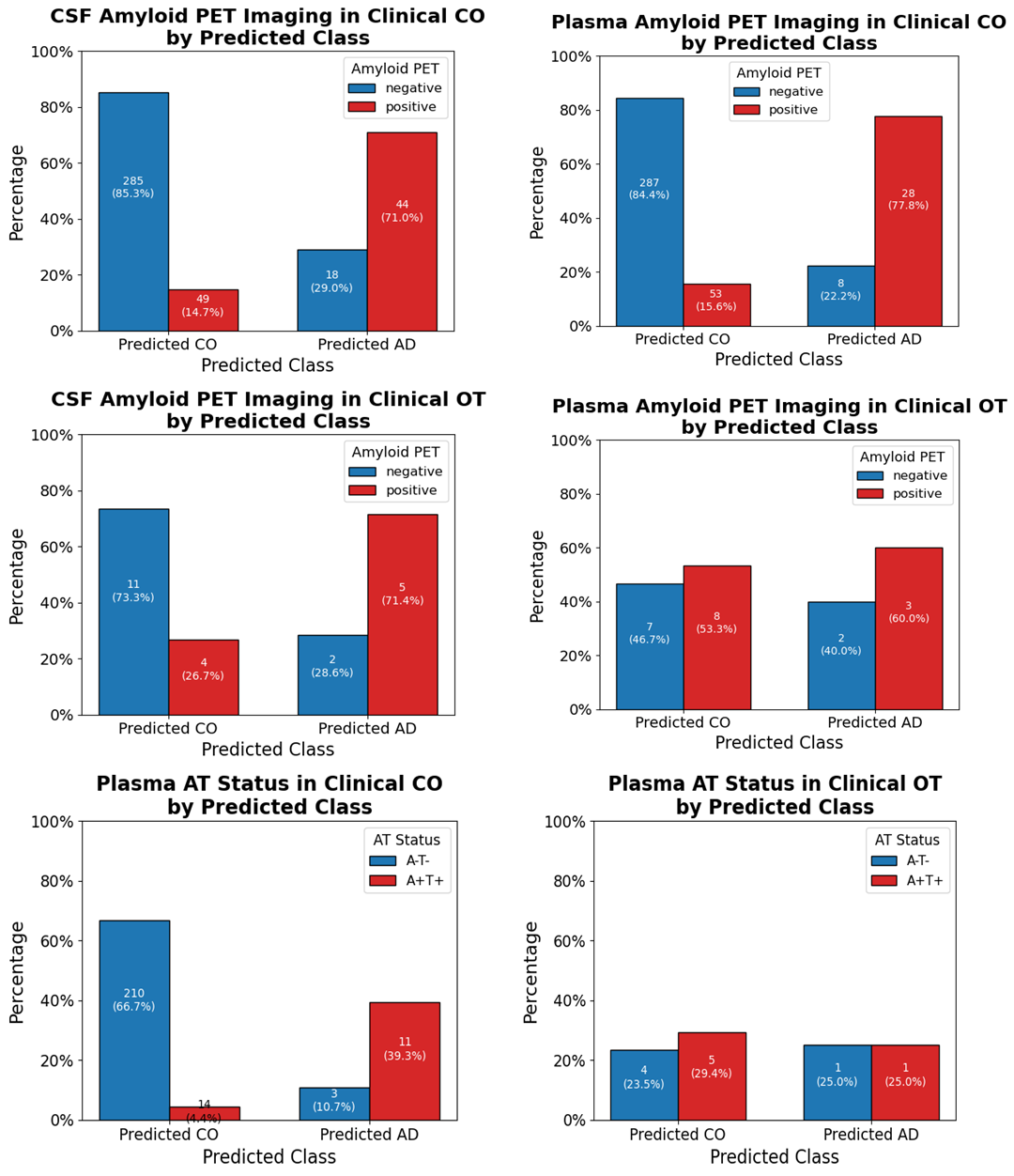

**Supplementary Figure 16. Established AD biomarkers corroborate AI-driven re-classification in clinically “cognitively unimpaired” (CO) and “other/indeterminate” (OT) participants. A,** barplots of amyloid PET positivity in samples predicted as CO compared to samples predicted as AD, among clinically-defined controls. Right, CSF; left, plasma. **B,** barplots of amyloid PET positivity in samples predicted as CO compared to samples predicted as AD, among individuals with evidence of cognitive decline but no conclusive diagnosis (OT). Right, CSF; left, plasma. **C,** barplots of CSF AT status (amyloid and tau negative, A-T-; amyloid and tau positive, A+T+) in samples predicted as CO compared to samples predicted as AD within plasma samples. Left, comparison in clinically-defined controls; right, comparison in individuals without a conclusive diagnosis (OT).

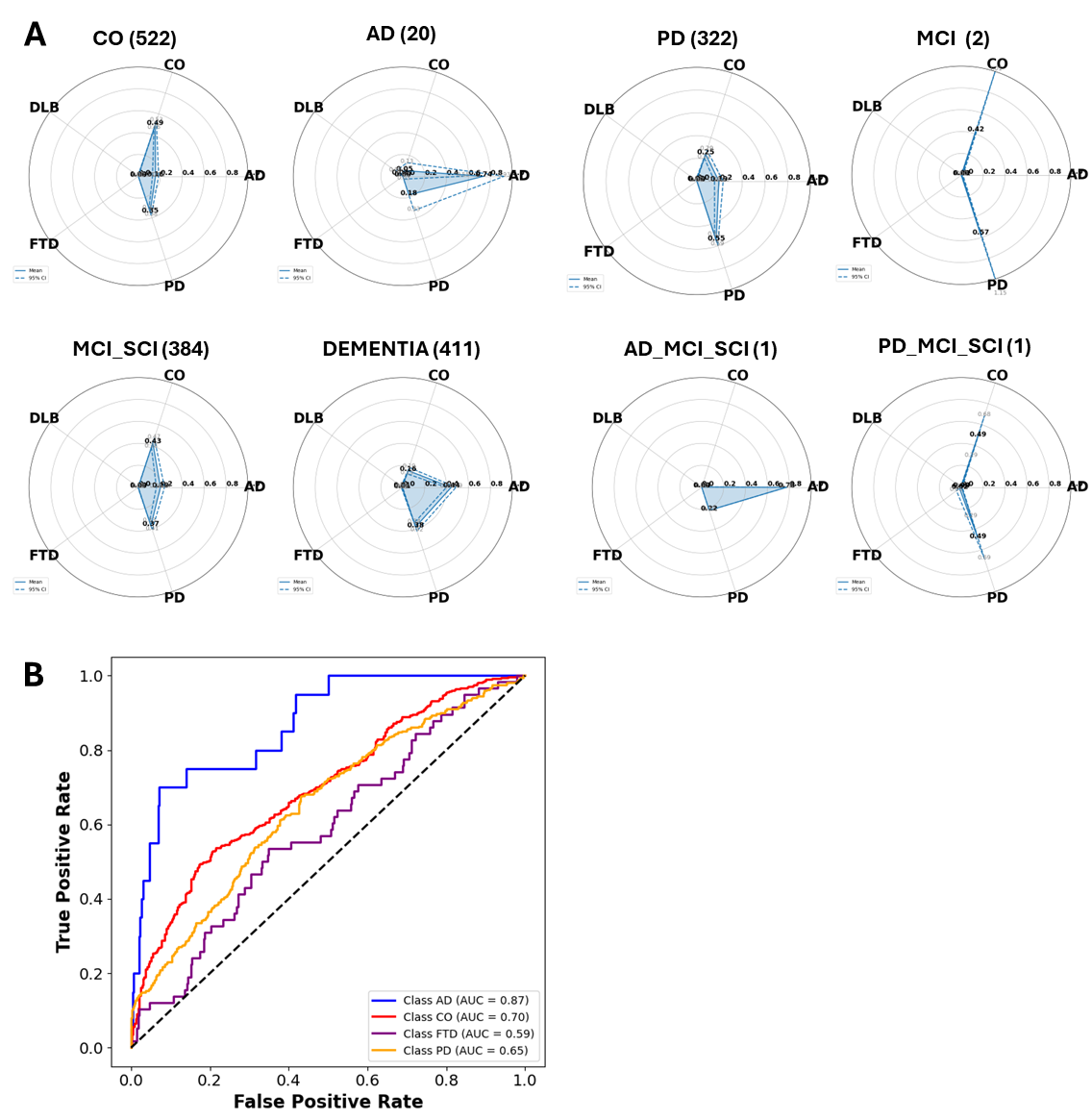
**Supplementary Figure 17. GNPC CSF Validation. A,** radar plots for different disease categories representing the mean probabilities of each class. Clockwise from top left, five-class probabilities for clinically-defined controls (CO), Alzheimer’s cases (AD), Parkinson’s cases (PD), mild cognitive impairment (MCI), PD & MCI (PD_MCI_SCI), AD & MCI (AD_MCI_SCI), evidence of cognitive impairment without clear diagnosis (DEMENTIA), and MCI/SCI (MCI_SCI). **B,** Area under the Receiver-Operator Curve (AUC) plot for CSF GNPC data. For each, AUC was calculated based on a one vs all paradigm.

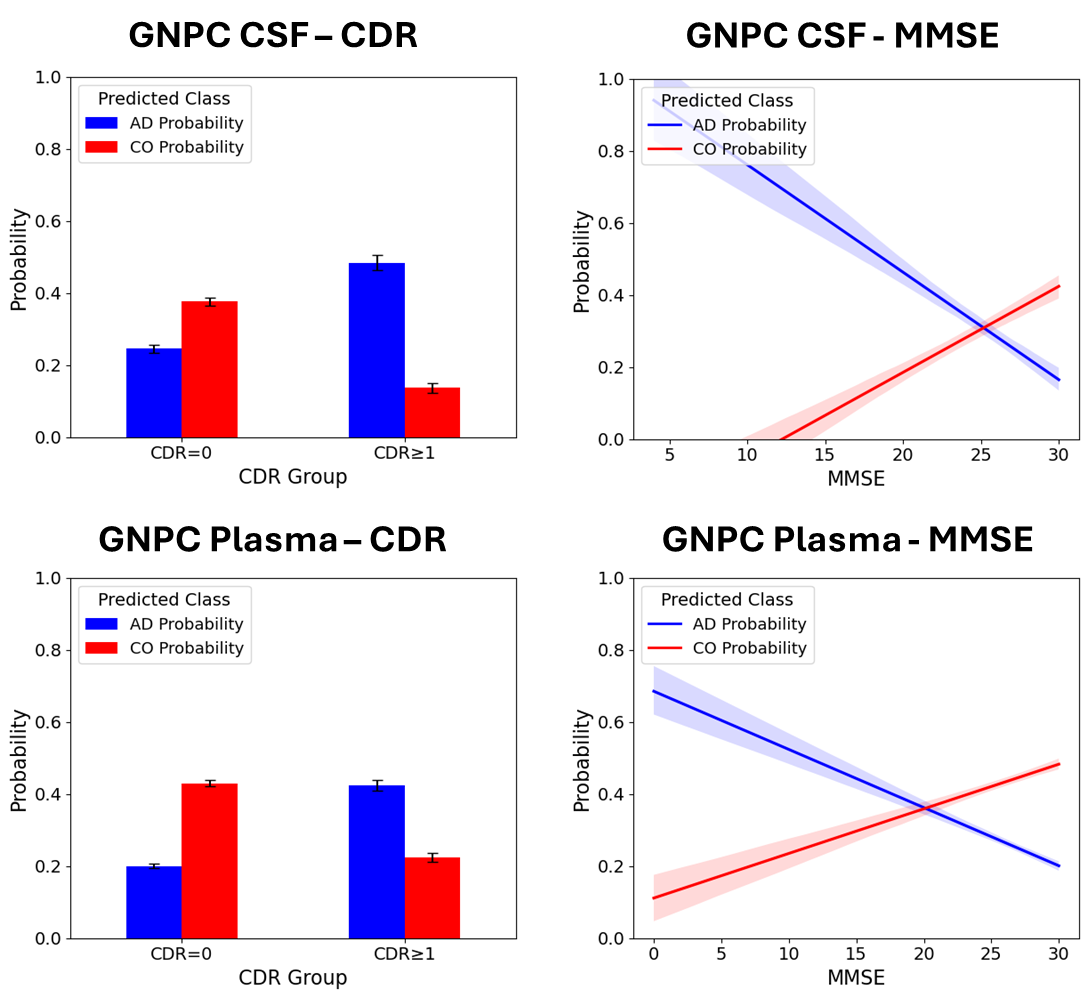
**Supplementary Figure 18. Predicted AD and CO class probabilities stratified by cognitive impairment severity.** Bar plots (left) and regression plots (right) illustrate predicted probabilities for AD (blue) and CO (red) classes in relation to clinical severity, as measured by Clinical Dementia Rating (CDR) groups (top: CSF-based model; bottom: plasma-based model) and Mini-Mental State Examination (MMSE) scores (right panels). In bar plots, individuals were dichotomized into cognitively normal (CDR=0) versus impaired (CDR≥1), and group means ± s.e.m. were shown. In regression plots, probability trajectories across the full MMSE range were modeled using non-parametric smoothing (Seaborn regplot) with 95% confidence intervals.

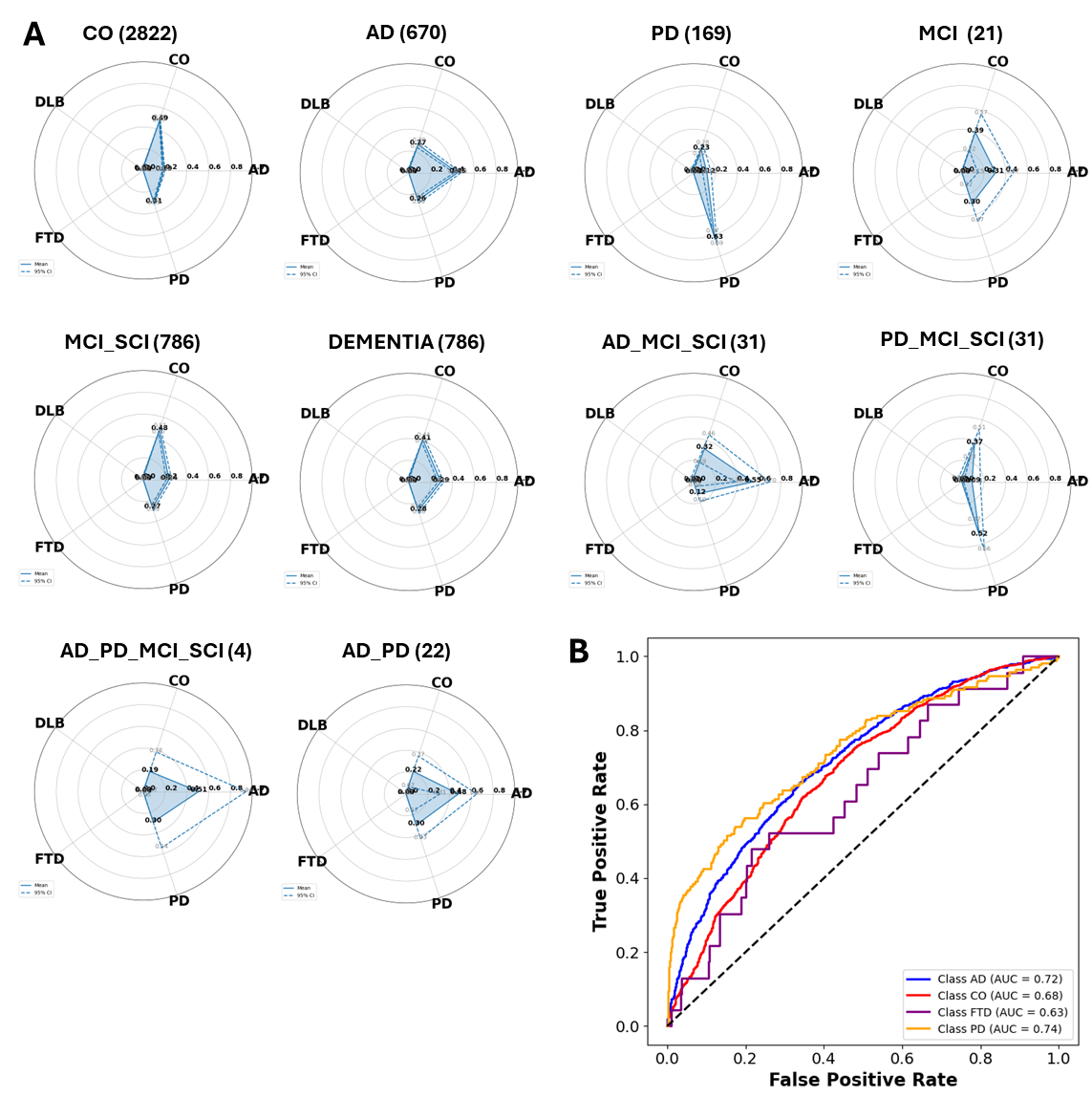
**Supplementary Figure 19. GNPC Plasma Validation. A,** radar plots for different disease categories representing the mean probabilities of each class. Row 1: five-class mean probabilities for clinically-defined controls (CO), Alzheimer’s cases (AD), Parkinson’s cases (PD), mild cognitive impairment (MCI). Row 2: mean probabilities for mild cognitive impairment/suggestive cognitive impairment (MCI_SCI), evidence of cognitive impairment without clear diagnosis (DEMENTIA), AD & MCI (AD_MCI_SCI), and PD & MCI (PD_MCI_SCI). Row 3: mean probabilities for AD, PD, & MCI (AD_PD_MCI_SCI) and AD & PD (AD_PD). **B,** Area under the Receiver-Operator Curve (AUC) plot for plasma GNPC data. For each, AUC was calculated based on a one vs all paradigm.
